# Rare coding variation illuminates the allelic architecture, risk genes, cellular expression patterns, and phenotypic context of autism

**DOI:** 10.1101/2021.12.20.21267194

**Authors:** Jack M. Fu, F. Kyle Satterstrom, Minshi Peng, Harrison Brand, Ryan L. Collins, Shan Dong, Lambertus Klei, Christine R. Stevens, Caroline Cusick, Mehrtash Babadi, Eric Banks, Brett Collins, Sheila Dodge, Stacey B. Gabriel, Laura Gauthier, Samuel K. Lee, Lindsay Liang, Alicia Ljungdahl, Behrang Mahjani, Laura Sloofman, Andrey Smirnov, Mafalda Barbosa, Alfredo Brusco, Brian H.Y. Chung, Michael L. Cuccaro, Enrico Domenici, Giovanni Battista Ferrero, Jay J. Gargus, Gail E. Herman, Irva Hertz-Picciotto, Patricia Maciel, Dara S. Manoach, Maria Rita Passos-Bueno, Antonio M. Persico, Alessandra Renieri, Flora Tassone, Elisabetta Trabetti, Gabriele Campos, Marcus C.Y. Chan, Chiara Fallerini, Elisa Giorgio, Ana Cristina Girard, Emily Hansen-Kiss, So Lun Lee, Carla Lintas, Yunin Ludena, Rachel Nguyen, Lisa Pavinato, Margaret Pericak-Vance, Isaac Pessah, Evelise Riberi, Rebecca Schmidt, Moyra Smith, Claudia I.C. Souza, Slavica Trajkova, Jaqueline Y.T. Wang, Mullin H.C. Yu, The Autism Sequencing Consoritum (ASC), Broad Institute Center for Common Disease Genomics (Broad-CCDG), iPSYCH-BROAD Consortium, David J. Cutler, Silvia De Rubeis, Joseph D. Buxbaum, Mark J. Daly, Bernie Devlin, Kathryn Roeder, Stephan J. Sanders, Michael E. Talkowski

**Affiliations:** Center for Genomic Medicine, Massachusetts General Hospital, Boston, MA 02114, USA; Program in Medical and Population Genetics, Broad Institute of MIT and Harvard, Cambridge, MA 02142, USA; Department of Neurology, Massachusetts General Hospital and Harvard Medical School, Boston, MA 02114, USA; Stanley Center for Psychiatric Research, Broad Institute of MIT and Harvard, Cambridge, MA 02142, USA; Analytic and Translational Genetics Unit, Department of Medicine, Massachusetts General Hospital, Boston, MA 02114, USA; Department of Statistics and Data Science, Carnegie Mellon University, Pittsburgh, PA 15213, USA; Pediatric Surgical Research Laboratories, Massachusetts General Hospital, Boston, MA 02114, USA; Program in Bioinformatics and Integrative Genomics, Harvard Medical School, Boston, MA 02115, USA; Department of Psychiatry, UCSF Weill Institute for Neurosciences, University of California San Francisco, San Francisco, CA 94143, USA; Department of Psychiatry, University of Pittsburgh School of Medicine, Pittsburgh, PA 15213, USA; Data Sciences Platform, The Broad Institute of MIT and Harvard,Cambridge, MA 02142, USA; Seaver Autism Center for Research and Treatment, Icahn School of Medicine at Mount Sinai, New York, NY 10029, USA; Department of Psychiatry, Icahn School of Medicine at Mount Sinai, New York, NY 10029, USA; The Mindich Child Health and Development Institute, Icahn School of Medicine at Mount Sinai, New York, NY 10029, USA; Genomics Platform, The Broad Institute of MIT and Harvard, Cambridge, MA 02142, USA; Department of Medical Epidemiology and Biostatistics, Karolinska Institutet, 171 77 Stockholm, Sweden; Department of Genetics and Genomic Sciences, Icahn School of Medicine at Mount Sinai, New York, NY 10029, USA; Department of Medical Sciences, University of Torino, Turin, 10126, Italy; Medical Genetics Unit, “Città della Salute e della Scienza” University Hospital, Turin, 10126, Italy; Department of Pediatrics & Adolescent Medicine, Duchess of Kent Children’s Hospital, The University of Hong Kong, Hong Kong Special Administrative Region 999077, China; The John P Hussman Institute for Human Genomics, The University of Miami Miller School of Medicine, Miami, FL 33136, USA; Department of Cellular, Computational and Integrative Biology, University of Trento, 38122 Trento, Italy; Department of Public Health and Pediatrics, University of Torino, Turin, 10126, Italy; Center for Autism Research and Translation, University of California Irvine, Irvine, CA 92697, USA; The Research Institute at Nationwide Children’s Hospital, Columbus, OH 43205, USA; MIND (Medical Investigation of Neurodevelopmental Disorders) Institute, University of California Davis, Davis, CA 95616, USA; Life and Health Sciences Research Institute, School of Medicine, University of Minho, Campus de Gualtar, 4710-057 Braga, Portugal; Department of Psychiatry, Massachusetts General Hospital and Harvard Medical School, Boston, MA 02114, USA; Centro de Pesquisas sobre o Genoma Humano e Células tronco, Instituto de Biociências, Universidade de São Paulo, São Paulo, 05508090, Brazil; Interdepartmental Program “Autism 0-90”, “Gaetano Martino” University Hospital, University of Messina, Messina I-98125, Italy; Med Biotech Hub and Competence Center, Department of Medical Biotechnologies, University of Siena, Ital; Medical Genetics, University of Siena, 53100 Siena, Italy; Genetica Medica, Azienda Ospedaliera Universitaria Senese, 53100 Siena, Italy; Department of Biochemistry and Molecular Medicine, University of California Davis, School of Medicine, Sacramento, CA 95817, USA; Department of Neurosciences, Biomedicine and Movement Sciences, Section of Biology and Genetics, University of Verona, 37134 Verona, Italy; Service for Neurodevelopmental Disorders, University Campus Bio-medico of Rome, 00128 Rome, Italy; Department of Molecular Biosciences, University of California Davis, School of Veterinary Medicine, Davis, CA 95616, USA; Department of Human Genetics, Emory University School of Medicine, Atlanta, GA 30322 USA; Friedman Brain Institute, Icahn School of Medicine at Mount Sinai, New York, NY 10029, USA; Department of Neuroscience, Icahn School of Medicine at Mount Sinai, New York, NY 10029, USA; Department of Genetics, Harvard Medical School, Boston, MA 02115, USA; Institute for Molecular Medicine Finland (FIMM), University of Helsinki, 00290 Helsinki, Finland; Computational Biology Department, Carnegie Mellon University, Pittsburgh, PA 15213, USA; Department of Psychiatry, Graduate School of Medicine, Nagoya University, Nagoya 466-8550, Japan; The Lundbeck Foundation Initiative for Integrative Psychiatric Research, iPSYCH, 8000 Aarhus, Denmark; Center for Genomics and Personalized Medicine, 8000 Aarhus, Denmark; Department of Biomedicine - Human Genetics, Aarhus University, 8000 Aarhus, Denmark; Bioinformatics Research Centre, Aarhus University, 8000 Aarhus, Denmark; Institute for Juvenile Research, Department of Psychiatry, University of Illinois at Chicago, Chicago, IL 60608, USA; Department of Internal Medicine, University of Utah, Salt Lake City, UT 84132, USA; Department of Psychiatry, Huntsman Mental Health Institute, University of Utah, Salt Lake City, UT 84108, USA; Grupo de Medicina Xenómica, Centro de Investigación en Red de Enfermedades Raras (CIBERER), CIMUS, Universidade de Santiago de Compostela, 15782 Santiago de Compostela, Spain; Neurogenetics group, Instituto de Investigación Sanitaria de Santiago (IDIS-SERGAS), 15706 Santiago de Compostela, Spain; Department of Child and Adolescent Psychiatry, Psychosomatics and Psychotherapy, Goethe University Frankfurt, 60528 Frankfurt, Germany; Center for Neonatal Screening, Department for Congenital Disorders, Statens Serum Institut, 2300 Copenhagen, Denmark; Department of Pediatrics, Icahn School of Medicine at Mount Sinai, New York, NY 10029, USA; Department of Public Health, Ben-Gurion University of the Negev, Beer-Sheva, 841050, Israel; National Autism Research Center of Israel, Ben-Gurion University of the Negev, 8410501, Beer-Sheva, Israel; Psychiatric & Neurodevelopmental Genetics Unit, Department of Psychiatry, Massachusetts General Hospital, Boston, MA 02114, USA; Department of Child and Adolescent Psychiatry, Hospital General Universitario Gregorio Marañón, IiSGM, CIBERSAM, School of Medicine Complutense University, 28009 Madrid, Spain; Department of Child Psychiatry, Tampere University and Tampere University Hospital, 33520 Tampere, Finland; Program in Genetics and Genome Biology, The Centre for Applied Genomics, The Hospital for Sick Children, Toronto, ON M5G 0A4, Canada; Department of Molecular Genetics and McLaughlin Centre, University of Toronto, Toronto, ON M5S, Canada; Harvard Medical School, Boston, MA 02115, USA; Fundación Pública Galega de Medicina Xenómica, Servicio Galego de Saúde (SERGAS), 15706 Santiago de Compostela, Spain; Division of Genetics and Genomics, Boston Children’s Hospital, Boston, MA 02115, USA; Medical Genomics Center, Nagoya University Hospital, Nagoya 466-8550, Japan; Department of Clinical Chemistry, Fimlab Laboratories and Finnish Cardiovascular Research Center-Tampere, Faculty of Medicine and Health Technology, Tampere University, 33520 Tampere, Finland; The Azrieli National Center for Autism and Neurodevelopment Research, Ben-Gurion University of the Negev, 8410501, Beer-Sheva, Israel; Pre-School Psychiatry Unit, Soroka University Medical Center, 8457108, Beer Sheva, Israel; Children’s Hospital of Philadelphia, Phladelphia, PA 19104; Department of Psychiatry, University of Utah, Salt Lake City, UT 84108, USA; Life Span Institute and Kansas Center for Autism Research and Training, University of Kansas, Lawrence, KS 66045; Institute for Glyco-core Research (iGCORE), Nagoya University, Furo-cho, Chikusa-ku, Nagoya 464-8601, Japan; Department of Environmental Medicine and Public Health, Icahn School of Medicine at Mount Sinai, New York, NY 10029, USA; Norwegian Institute of Public Health, Oslo, 0213, Norway; Department of Psychiatry, University of Cincinnati, Cincinnati OH 45219 USA; Department of Epidemiology, Harvard T.H. Chan School of Public Health, 655 Huntington Avenue, Building II 2nd Floor, Boston, MA 02115, USA; Metabolomics Platform, Broad Institute of Harvard and MIT, 415 Main St, Cambridge, MA 02142, USA; The Broad Institute of MIT and Harvard, Cambridge, MA 02142, USA; Verve Therapeutics, Cambridge, MA 02139, USA

## Abstract

Individuals with autism spectrum disorder (ASD) or related neurodevelopmental disorders (NDDs) often carry disruptive mutations in genes that are depleted of functional variation in the broader population. We build upon this observation and exome sequencing from 154,842 individuals to explore the allelic diversity of rare protein-coding variation contributing risk for ASD and related NDDs. Using an integrative statistical model, we jointly analyzed rare protein-truncating variants (PTVs), damaging missense variants, and copy number variants (CNVs) derived from exome sequencing of 63,237 individuals from ASD cohorts. We discovered 71 genes associated with ASD at a false discovery rate (FDR) ≤ 0.001, a threshold approximately equivalent to exome-wide significance, and 183 genes at FDR ≤ 0.05. Associations were predominantly driven by *de novo* PTVs, damaging missense variants, and CNVs: 57.4%, 21.2%, and 8.32% of evidence, respectively. Though fewer in number, CNVs conferred greater relative risk than PTVs, and repeat-mediated *de novo* CNVs exhibited strong maternal bias in parent-of-origin (e.g., 92.3% of 16p11.2 CNVs), whereas all other CNVs showed a paternal bias. To explore how genes associated with ASD and NDD overlap or differ, we analyzed our ASD cohort alongside a developmental delay (DD) cohort from the deciphering developmental disorders study (DDD; n=91,605 samples). We first reanalyzed the DDD dataset using the same models as the ASD cohorts, then performed joint analyses of both cohorts and identified 373 genes contributing to NDD risk at FDR ≤ 0.001 and 662 NDD risk genes at FDR ≤ 0.05. Of these NDD risk genes, 54 genes (125 genes at FDR ≤ 0.05) were unique to the joint analyses and not significant in either cohort alone. Our results confirm overlap of most ASD and DD risk genes, although many differ significantly in frequency of mutation. Analyses of single-cell transcriptome datasets showed that genes associated predominantly with DD were strongly enriched for earlier neurodevelopmental cell types, whereas genes displaying stronger evidence for association in ASD cohorts were more enriched for maturing neurons. The ASD risk genes were also enriched for genes associated with schizophrenia from a separate rare coding variant analysis of 121,570 individuals, emphasizing that these neuropsychiatric disorders share common pathways to risk.

## Background

Autism spectrum disorder (ASD) affects over 1.7% of children in the United States^1^. Epidemiological studies have repeatedly demonstrated that ASD is highly heritable^2^, with the majority of risk stemming from common genetic variants, each of small effect, acting additively across the genome^3,4^. However, in at least 10% of cases, rare and *de novo* genetic variants confer substantial risk^5–9^. Exome sequencing provides an efficient method to detect these rare and *de novo* variants, which has led to the association of numerous genes with ASD through repeated observations of such variants across independent cases^10^.

Beyond ASD, exome sequencing has enabled the discovery of genes associated with overlapping and distinct genetic architectures across a spectrum of developmental and neuropsychiatric disorders^6,10–14^. These exome studies have largely focused on analyses of single nucleotide variants (SNVs) and insertion/deletion variants (indels), in particular *de novo* protein-truncating variants (PTVs) and missense variants, with several studies noting modest enrichment of rare inherited variants as well^10,15^. The relative enrichment of *de novo* PTVs in cases varies by ascertainment strategy: burden is greatest in individuals with developmental delay (DD), intellectual disability, or multi-system congenital anomalies; moderate in individuals with ASD or isolated developmental anomalies; and lowest in schizophrenia and other neuropsychiatric disorders^10,11,13,14,16,17^. Indeed, hundreds of genes have now been discovered across this spectrum of developmental disorders, with associations driven by phenotype severity and cohort size^18–20^.

Furthermore, it has been well established that ASD and DD cases harbor an excess of very large copy number variants (CNVs) compared to unaffected siblings^21–27^. While these CNVs of large genomic segments represent an approximately 3.5-fold increase in ASD risk^28^, their incorporation into genomic studies has long represented a significant technical challenge. Early studies using microarrays were limited to CNVs of hundreds of kilobases to megabases^12,21,25,29^. Among the most significant findings from these studies were recurrent genomic disorder (GD) loci associated with syndromic phenotypes that arose due to mispairing of long homologous segments, a mechanism known as non-allelic homologous recombination (NAHR)^12,25,28,30^. Due to their high mutation rate, these reciprocal GD regions collectively represent a significant source of genetic risk for neurodevelopmental disorders (NDDs)^12,25,28,30^.

What remains less clear is the relative burden of CNVs at the resolution of individual genes or exons in ASD etiology, as studies to date have been limited by the relatively small samples assessed by whole-genome sequencing (WGS) and the high false positive rates for existing exome-based CNV discovery methods^31^. Prior studies by the Exome Aggregation Consortium (ExAC^32^), 1000 Genomes Project^33–35^, and Genome Aggregation Database (gnomAD^36^) have established size distributions and mutation rates for structural variants (SVs) across human populations, while studies leveraging long-read WGS have provided further insights into the broader swath of human SVs, over 99% of which are not readily detectable by microarray^37,38^. Here, we demonstrate that significant improvements in computational models to normalize exome sequencing read depth, combined with appropriate quality control filters, enable discovery of even small rare coding CNVs with validation rates comparable to indels. These methods allow us to build upon prior studies^22^ to now jointly analyze rare coding SNVs, indels, and CNVs at the resolution of individual genes and exons from large-scale ASD datasets.

Rare variant discovery can also be enhanced through the integration of functional data, particularly in cohorts such as ASD that display strong selective pressure and reduced fecundity^10^. One such measure is the recently developed ‘loss-of-function observed/expected upper bound fraction’ (LOEUF) score^39^, which is a continuous measure of selective pressure against PTVs in each gene. Similarly, the ‘missense badness, PolyPhen-2, and constraint’ (MPC) score^40^ is one of several measures of the estimated deleteriousness of missense variation. In this study, we used a Bayesian statistical framework, the Transmission and *De Novo* Association (TADA) model^41^, to incorporate these functional annotations and jointly analyze coding SNVs, indels, and CNVs across the largest exome-sequenced ASD and DD cohorts to date, comprising 63,237 individuals from ASD cohorts (20,627 ASD-affected individuals) and 91,605 samples from DD cohorts (31,058 DD-affected individuals). Our analyses identify hundreds of genes associated with these disorders and reveal significant overlap, as well as substantial heterogeneity, in the genes associated with each phenotype and in the cell types in which they are enriched during early neuronal development. Overall, these analyses provide new insights into the contributions of rare coding variation in NDDs, including broad overlap and nuanced distinctions of genetic risk and its influence on specific pathways and developmental trajectories.

## RESULTS

### Patterns of coding variants in a large ASD exome dataset

We aggregated whole exome sequencing (WES) data across 33 ASD research cohorts, totalling 63,237 individuals: 15,036 affected offspring, 5,492 unaffected offspring, 28,522 parents, as well as 5,591 affected and 8,597 unaffected individuals from case-control studies (one sample exists in our dataset as both mother and affected offspring, **Fig. 1a, Supplementary Tables 1, 2, 3, 4.1**). Data from 35% of these individuals had not previously been analyzed or published. All accessible DNA sequence reads were aligned to the GRCh38 human reference genome and coding SNVs, indels, and CNVs were identified using GATK haplotype caller^42^ while a new method, GATK-gCNV^43^, was used for CNV delineation. Variant counts were consistent across cohorts, with an average of 1.64 (1.66/affected, 1.57/unaffected) *de novo* SNVs, 0.18 (0.18/affected, 0.16/unaffected) *de novo* indels, and 0.035 (0.042/affected, 0.014/unaffected) *de novo* CNVs per offspring.

**Fig. 1.**
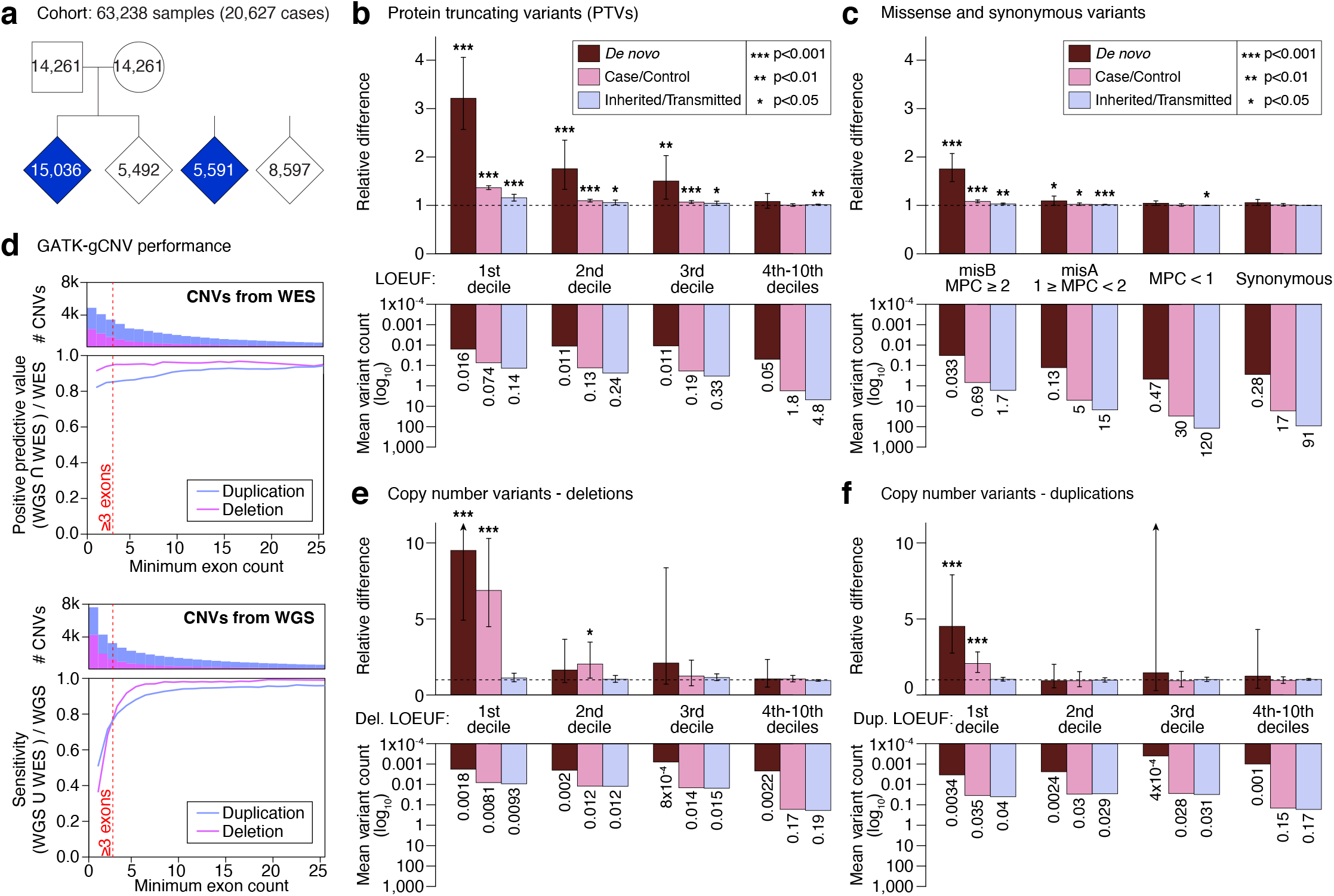
Overview of SNV/indel and CNV rates in ASD by mode of inheritance. a, The ASD cohort for SNV/indel analysis consisted of 49,049 family-based samples (15,036 cases) and 14,188 case/control samples (5,591 cases). b, The relative difference in PTV frequency between cases and unaffected controls (top) and average per-sample variant count in unaffected controls (bottom) across inheritance classes (color) and LOEUF deciles (5,446 genes in top 3 deciles of LOEUF). Using a binomial test, we found significant enrichment in cases for PTVs among the most constrained genes (lower LOEUF deciles), which weakened as negative selection against PTVs is relaxed (higher LOEUF deciles). c, Equivalent information and statistical significance for missense and synonymous variants. Synonymous variants were not enriched in cases or controls, as evaluated via binomial tests, in keeping with well-calibrated variant calling. d, The GATK-gCNV exome CNV detection pipeline when compared against WGS on overlapping samples achieved a sensitivity of 82% and positive predictive value of 89% for rare CNVs (<1% site frequency) that spanned 3 or more callable exons (red line). e, The relative difference in variant frequency between cases and controls for deletions. Using binomial tests, we found that deletions overlapping genes in the lowest LOEUF decile were observed at an elevated rate in affected individuals. f, Equivalent analysis for duplications, demonstrating a similar pattern of enrichment compared to deletions but with a weaker effect. Abbreviations: PTV: protein truncating variant; CNV: copy number variant; WGS: whole genome sequencing; WES: whole exome sequencing; misB: missense variants with MPC score >=2; misA: missense variants with MPC score >=1 and <2; Del: Deletion CNV; Dup: Duplication CNV. Statistical tests: b,c,e,f: binomial test.

Consistent with prior studies, we observed more PTVs and damaging missense variants in individuals with ASD compared to unaffected individuals (**Fig. 1b-c**). Enrichment of *de novo* and inherited PTVs in ASD cases was greatest in genes with the strongest evidence of selective constraint, represented by low LOEUF scores^39^ (**Supplementary Tables 4.2-4.7**). Both *de novo* and inherited PTVs were enriched in the top three deciles of LOEUF (binomial test; 5,446 total genes; **Fig. 1b**). We grouped missense variants into three bins based on MPC score, which we termed MisB (MPC ≥ 2) and MisA (2 > MPC ≥ 1), with MPC < 1 for all remaining missense variants. MisB variants and, to a lesser degree, MisA variants were significantly enriched in ASD individuals (**Fig. 1c**). Our aggregated cohort demonstrated the greatest risk from *de novo* variation, with modest but significant ASD risk observed in rare case/control (for which *de novo* status cannot be determined) and inherited PTVs in the top three deciles of constrained genes, as well as in MisB and MisA variants.

### Detection and analysis of rare and de novo CNVs in ASD

Previous studies using chromosomal microarrays have established a clear etiological role for large, rare and *de novo* CNVs in ASD cases compared to their unaffected siblings and the general population^21,22,25–27,44–46^. Despite their considerable impact on ASD risk, CNV discovery at the resolution of individual exons from exome sequencing has represented a significant technical challenge. To overcome these challenges, we applied and benchmarked a pipeline built around GATK-gCNV, a novel Bayesian read depth-based CNV discovery tool for short-read sequencing data^42^. Our pipeline includes a PCA-based method for inferring large systematic differences, such as different exome capture kits (**Supplementary Fig. 1**), and we performed extensive benchmarking of this method from 7,165 samples from ASD quartet families for which matching gold-standard CNV calls from microarray and WGS were available^47,48^. We measured sensitivity and positive predictive value (PPV) as a function of the number of captured exons from canonical protein-coding transcripts. Our application of GATK-gCNV achieved a sensitivity of 82% and a PPV of 89% for rare CNVs (site frequency < 1%) that spanned 3 or more such exons (**Fig. 1d, Supplementary Fig. 2**). When we considered *de novo* CNVs detected by WGS, GATK-gCNV achieved 82% sensitivity and 97.5% PPV for *de novo* CNV discovery at three-exon or greater resolution (**Supplementary Fig. 3**).

We applied GATK-gCNV to the 57,294 samples from the ASD cohort for which raw sequencing data were available (**Supplementary Methods, Supplementary Table 3**), including 13,694 affected and 5,007 unaffected offspring with data from both parents and 608 cases and 11,312 controls from non-family based cohorts. We focused our analysis on high quality CNVs spanning at least three captured protein-coding exons (Supplementary Methods), resulting in 16,889 rare (frequency < 1%) and 650 *de novo* autosomal CNVs, corresponding to an average of 0.90 (0.91/affected, 0.89/unaffected) rare and 0.035 (0.042/ affected, 0.014/unaffected) *de novo* CNVs per offspring.

Consistent with *de novo* coding CNVs mediating substantial risk for ASD, we observed that 3.93% of ASD cases and 1.40% of unaffected siblings harbored at least one *de novo* coding CNV, respectively (odds ratio [OR]: 2.88, p=9.6 × 10^−21^, Fisher’s exact). Splitting the CNVs by dosage (deletions versus duplications) and assigning each event to a LOEUF decile based on the most constrained gene within its boundaries, we observed that *de novo* deletions with at least one PTV intolerant gene showed the greatest degree of enrichment in ASD cases across all variant classes evaluated (OR: 9.10, p=2.81×10^−20^, binomial test), with a relative difference approximately three-fold higher than that observed for *de novo* PTVs in the same constraint decile (p=2.9×10^−4^, simulation test). Enrichment of deletions was also observed in case/ control but not in rare inherited analyses. A similar but less significant pattern was observed for duplications across constraint deciles and modes of inheritance (**Fig. 1e-f**). Additionally, duplications with breakpoints residing within a gene have been suggested to harbor risk differently than duplications of an entire gene, likely through a loss of function mechanism^28,36^. Leveraging our *de novo* duplication data, we found that partially duplicated genes in cases harbored more *de novo* PTV evidence compared to partially duplicated genes in controls (1.3 fold, p=1.7×10^−3^, binomial test).

To characterize the impact of exon and gene-level resolution in CNV discovery, we first considered 75 large genomic segments (**Supplementary Table 4.8**) associated with NDDs from curated literature^28,49–54^. Of the 650 *de novo* CNVs discovered from WES here, 223 (34.3%) matched one of these loci with at least 50% reciprocal overlap, including 109 *de novo* GD deletions in cases compared to 6 in controls (OR: 6.7, p=3.9×10^−9^, Fisher’s exact) and 100 *de novo* GD duplications in cases compared to 8 in controls (OR: 4.6, p=5.4×10^−7^, Fisher’s exact, **Fig. 2b**). Considering rare inherited CNVs, we observed a non-significant trend towards enrichment of GD CNVs in cases (OR:1.3, p=0.07, Fisher’s exact). Excluding the GD loci, the remaining 427 *de novo* CNVs were also enriched in cases but with more modest effect sizes (*de novo* non-GD deletion: OR = 2.8, p=1.5×10^−8^, Fisher’s exact; *de novo* non-GD duplications: OR = 1.8, p=5.1×10^−3^, Fisher’s exact). A greater degree of enrichment was observed for non-GD deletions that overlapped a constrained gene (OR: 5.10, p=9.7×10^−11^, Fisher’s exact, **Fig. 2c**) than for those that did not (OR: 1.32, p=0.32, Fisher’s exact, Fig. 2c) or for *de novo* PTVs in constrained genes (OR: 2.74, p=8.5×10^−34^, Fisher’s exact, **Fig. 2c**). Effect sizes for *de novo* duplications were consistently smaller, although similar patterns were observed in relation to GD loci and constrained genes (**Fig. 2c**). Finally, when we considered *de novo*, non-GD CNVs that altered one of 71 genes associated with ASD risk (FDR ≤ 0.001, see below), we observed 46 variants in cases (29 deletions, 17 duplications) compared to none in siblings (p=8.5×10^−7^, Fisher’s exact). Considering the list of 373 genes implicated in both ASD and DD risk (see below) this increased to 130 *de novo* CNVs in cases and one in siblings (OR: 48.0, p=1.13×10^−16^, Fisher’s exact).

**Fig. 2.**
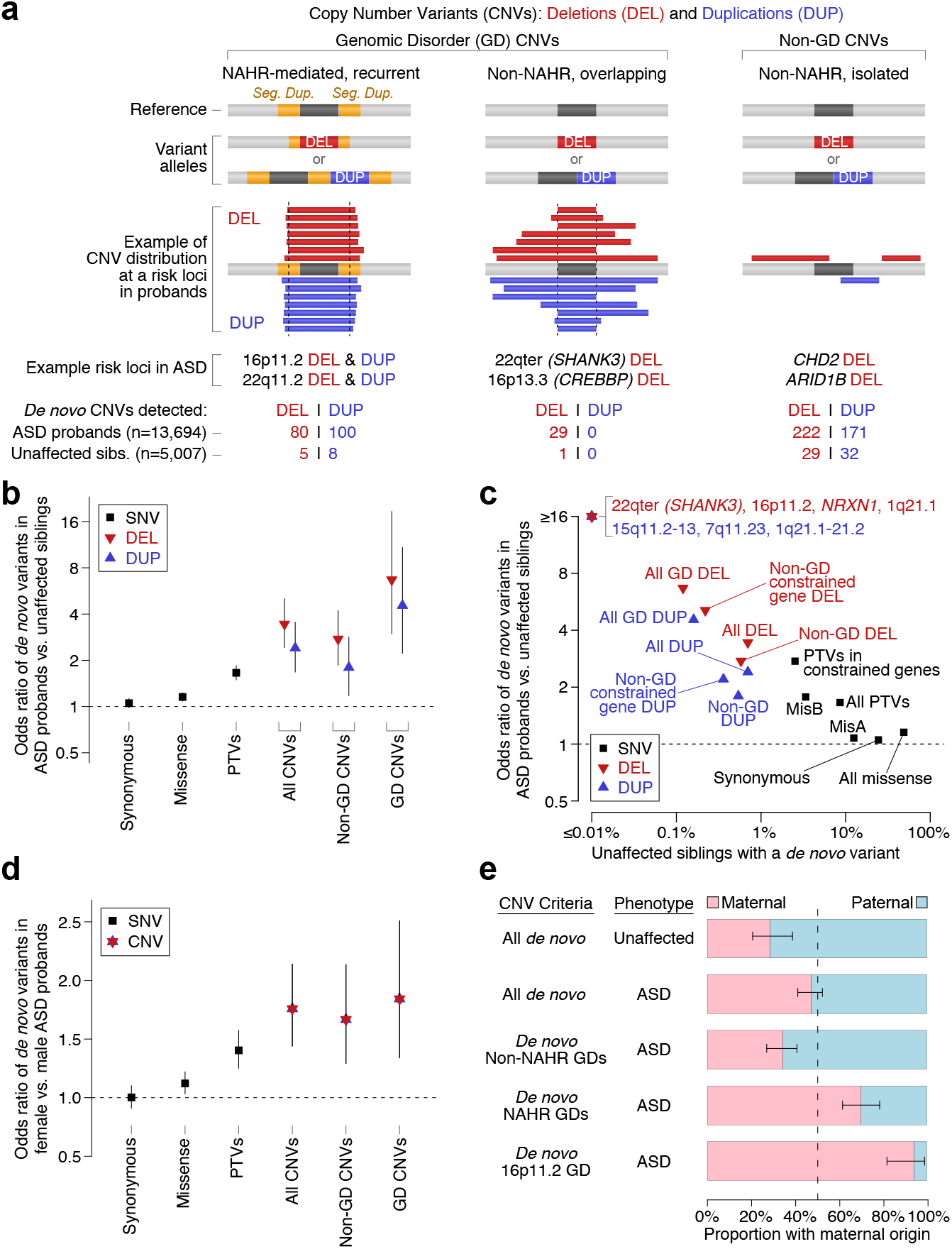
Contribution of CNVs to ASD by mechanism and genomic location. a, CNVs involve deletions (DEL) or duplications (DUP) of genomic segments, and include a subset of sites known as genomic disorder (GD) loci that reoccur in human disorders. GDs that are mediated on both ends by non-allelic homologous recombination (NAHR) have recurrent breakpoints in the population, whereas non-NAHR GDs are located at sites susceptible to rearrangements. CNVs outside of known GD loci can still contribute to ASD risk as more unique events. b, Using Fisher’s exact tests, we find that de novo CNVs are more frequent in affected than unaffected offspring, and show a higher elevation than that observed in de novo PTVs (p=1.3×10^−5^, logistic regression). c, High ORs are observed for GD CNVs, a subset of which have no events observed in controls in this cohort (e.g., 16p11.2 deletions, 15q11.2-q13 duplications). Excluding GD CNVs, having a CNV that overlaps a constrained gene (LOEUF < 0.4) confers greater risk than having CNVs that overlap only unconstrained genes (p=3.3×10^−4^, logistic regression). d, Using Fisher’s exact tests, we find that de novo risk-mediating variants are observed at a higher frequency in affected females than affected males. e, Parent-of-origin analysis of de novo CNVs using binomial tests shows maternal bias for NAHR-mediated CNVs at GD regions, which is especially pronounced for 16p11.2. Abbreviations: CNV: copy number variant; DEL: deletion CNV; DUP: duplication CNV; NAHR: non-allelic homologous recombination; PTV: protein truncating variant; GD: genomic disorder. Statistical tests: b,c,d: Fisher’s exact test, logistic regression; e: binomial test.

Consistent with a female protective effect in ASD, which proposes that a higher burden of risk factors is required for an ASD diagnosis in females^27,55^, the burden of *de novo* CNVs overall was higher in female cases than male cases (5.94% vs. 3.47%, OR:1.76, p=3.4×10^−8^, Fisher’s exact). This effect was more pronounced for CNVs than PTVs (p=2.6×10^−2^, logistic regression) or missense variants (p=2.0×10^−5^, logistic regression, **Fig. 2d**). While *de novo* SNVs and indels frequently arise on the paternal allele^9,44,56^, a maternal allele bias has been observed from *de novo* CNVs in ASD^57^. Using parental SNP data, we estimated the allelic parent-of-origin for 330 *de novo* CNVs (**Supplementary Methods**) and observed no overall bias (**Fig. 2e**, 130 maternal vs. 144 paternal, p=0.43, binomial test). However, restricting analyses to *de novo* CNVs at NAHR-mediated GD loci, we recapitulated prior findings with 70% arising on the maternal allele (70 maternal vs. 30 paternal, p = 7.9×10^−5^, binomial test). Including 20 additional samples from the Simons Searchlight project^58^, we find that 94.3% of 16p11.2 CNVs occur on the maternal allele (**Fig. 2e**, 33 maternal vs. 2 paternal, p=3.7×10^−8^, binomial test). In contrast, CNVs at non-NAHR-mediated GD loci showed a 65.5% paternal bias (**Fig. 2e**, 60 maternal vs. 114 paternal, p=5.2×10^−5^, binomial test), consistent with a mechanistic bias in CNV formation and previous findings of a paternal origin across all non-recurrent/non-pathogenic CNVs^48^.

### Integrated discovery of ASD-associated genes across variant types and inheritance classes

Our analyses identified enrichment of rare protein-coding variants in ASD cases with effect sizes varying by mode of inheritance, variant class (PTV, misB, misA, deletion, duplication), and evolutionary constraint. We sought to utilise these insights and the size of our cohort (63,237 samples) to refine gene discovery in ASD by extending the TADA analytic framework^6,10,22,41^ to include: (1) rare and *de novo* CNVs, (2) variants present in unaffected offspring, and (3) a continuous measure of evolutionary constraint from gnomAD (LOEUF^39^, **Supplementary Methods, Supplementary Fig. 4**). A Bayes Factor (BF) was calculated to represent evidence of association for each autosomal protein-coding gene across variant types and modes of inheritance, taking into account null mutation rates and prior relative risks, when appropriate (**Fig. 3**).

**Fig. 3.**
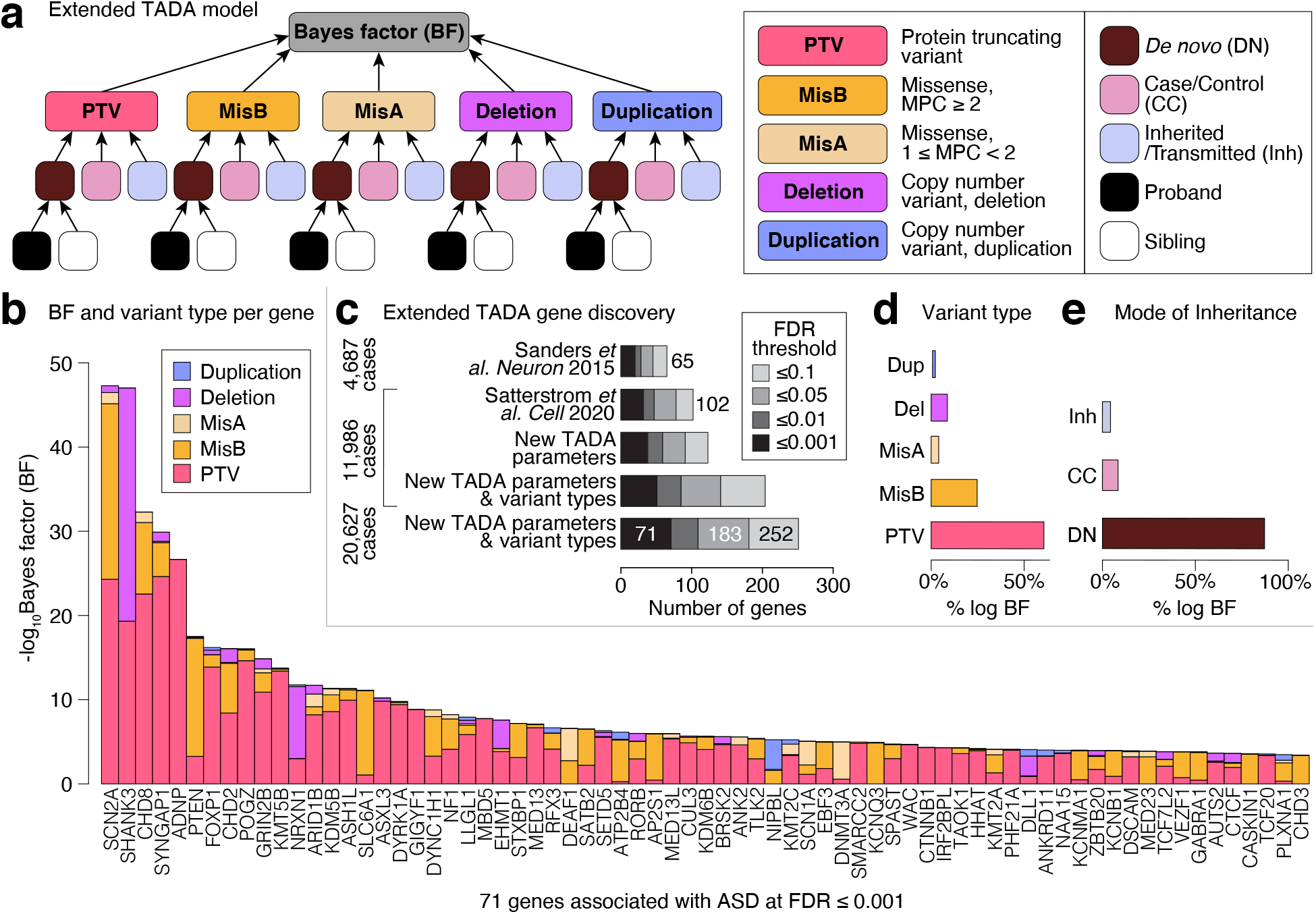
Integrating variant types and inheritance classes significantly boosts association power and reveals mutational biases. a, Our implementation of the TADA model includes de novo, case/control, and rare inherited modules for each variant type: PTV, MisB, MisA, deletion, and duplication. We leverage information from ASD probands as well as unaffected siblings in evaluating the effect of de novo variants. b, The evidence of ASD association contributed by each variant type for each of the 71 ASD genes with FDR ≤ 0.001. Some genes are predominantly associated through missense variants and duplications (e.g., PTEN, SLC6A1), suggesting mechanisms such as gain-of-function in contrast to haploinsufficient loss-of-function for genes with large contributions from PTVs and deletions. c, Applying TADA to our aggregated ASD dataset yields 71 genes at FDR ≤ 0.001, compared to 32 and 19 genes at the same threshold in previous studies on a subset of the samples (Satterstrom et al. 2020 and Sanders et al. 2015, respectively). Our expanded TADA model makes more efficient use of the available evidence of association by integrating more information into our statistical modeling. d, Across 71 ASD genes, the majority of evidence for ASD association is derived from PTVs and MisB variants and, e, de novo variants. Abbreviations: BF: Bayes factor; PTV: protein truncating variant; misB: missense variants with MPC score >=2; misA: missense variants with MPC score >=1 and <2; Del: deletion CNV; Dup: duplication CNV; Inh: inherited; CC: case/control; DN: de novo. Statistical tests: b Extended TADA model.

Applying this model to the aggregated ASD data (TADA-ASD) we identify 71 genes associated with ASD at a FDR ≤ 0.001 and 183 genes associated at FDR ≤ 0.05 (**Supplementary Table 4.9**). Notably, the FDR ≤ 0.001 threshold is approximately equivalent to Bonferroni significance when back-calculating a p-value and correcting for 18,128 autosomal genes (p < 2.8×10^−6^), making it comparable to other recent studies of schizophrenia^13^ and DD^11^. We focus on these 71 genes here, and illustrate in **Fig. 3** that 93% of these genes have contributions from multiple variant classes (**Fig. 3b**). They also represent a meaningful increase in ASD discovery -a prior study from Satterstrom *et al*. included a subset of 11,986 of the cases aggregated here, producing 32 ASD risk genes at this same FDR ≤ 0.001 threshold using the TADA parameters applied in that study, which increases to 51 genes when incorporating the updated model parameters and the CNVs discovered here (**Fig. 3c**). The BFs within TADA allow us to assess genomic architecture through the relative contributions across variant types and modes of inheritance (**Fig. 3d-e, Fig. 4a-b**). In keeping with haploinsufficiency as the primary pathogenic mechanism, PTVs and deletions account for over 90% of the evidence in 20 of the 71 ASD-associated genes (28.2%). However, for 10 genes (14.1%), over 90% of the evidence comes from missense variants and duplications (e.g., *DEAF1, SLC6A1*; **Fig. 4a**). Included within these 10 genes is *PLXNA1*, for which the evidence comes primarily from inherited missense variation localized within the Plexin domain of the encoded protein (**Fig. 4b-c**).

**Fig. 4.**
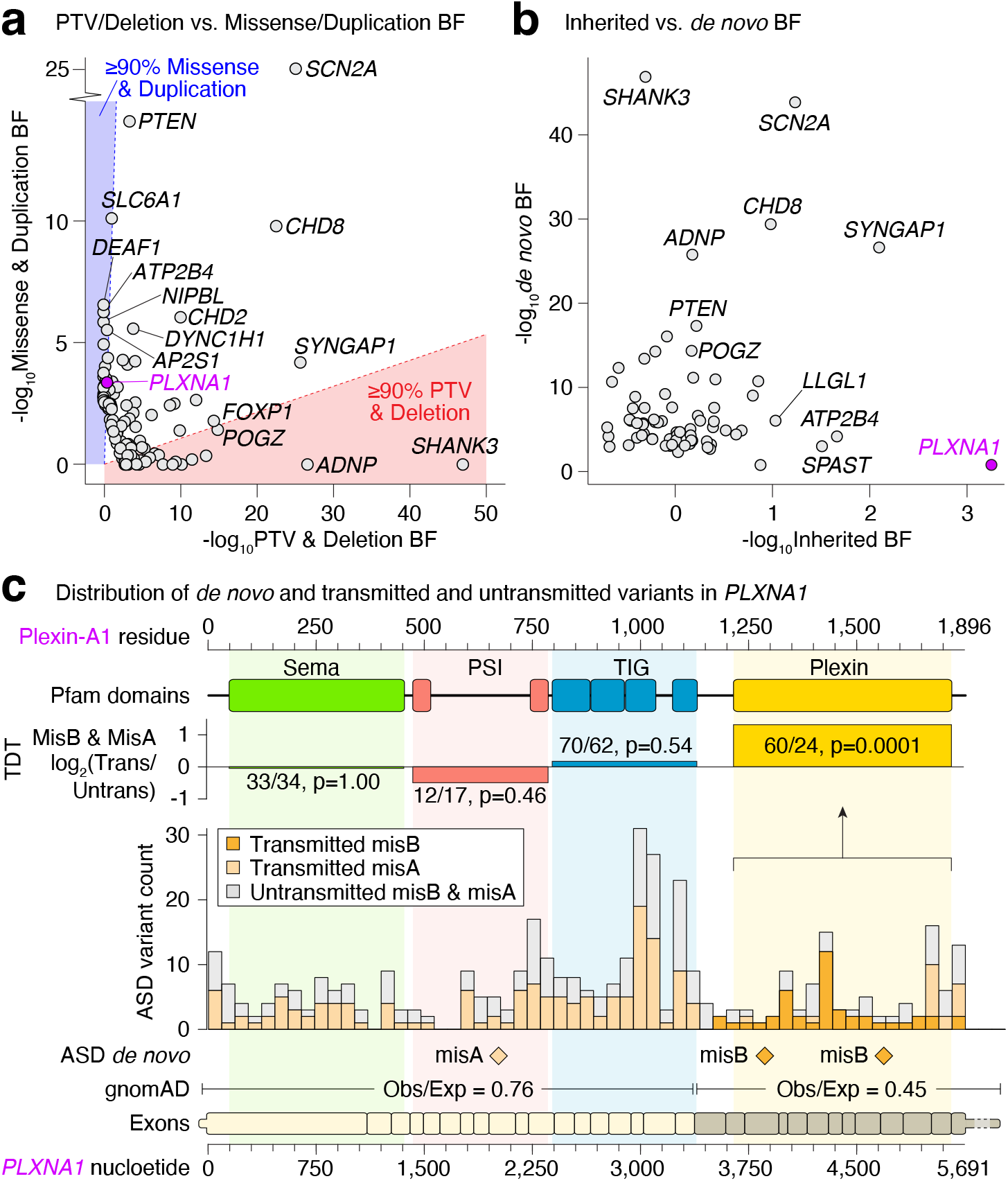
Relative contribution of evidence types in ASD risk genes. a, For 71 ASD-associated genes (FDR ≤ 0.001) the relative evidence of ASD association in the extended TADA model (log10BF) is shown for variants acting via a likely loss-of-function (LOF) mechanism (PTVs and deletions) on the x-axis versus variants that may act via alternative mechanisms (missense variants and duplications) on the y-axis. b, Equivalent plot of the relative evidence from de novo (x-axis) versus inherited (y-axis) variation for the same 71 ASD-associated genes. c, Evidence for ASD-association for the gene PLXNA1 is derived from de novo and transmitted missense variants, especially in the Plexin domain at the C-terminus of the Plexin-A1 protein. Abbreviations: BF: Bayes factor. Statistical tests: c: Transmission disequilibrium test.

We next sought to estimate the magnitude of the ASD risk imparted by the genes discovered here and described in **Fig. 3**. Within the 71 genes, *de novo* PTVs or damaging missense variants were detected in 4.0% of cases and 0.5% of controls, giving a combined OR of 8.19. Using cross validation (**Supplementary methods**), we refined this estimate by variant type and mode of inheritance with OR estimates ranging from 16.0 for *de novo* PTVs to 1.02 for inherited MisA (**Supplementary Table 4.10**). We also applied this cross validation approach to estimate the yield of future gene discovery efforts, predicting a greater than linear increase in gene discovery at this same statistical threshold from a two-fold increase in the current sample size (150 genes; 95% CI: 108-278) or a three-fold increase (344 genes; 95% CI: 218-500).

### Comparing the genetic architectures of ASD and general NDDs

ASD is frequently comorbid with other syndromic and non-syndromic NDDs, including intellectual disability (ID), and neurological disorders, including seizures^59,60^. Prior analyses have clearly demonstrated both overlapping and distinct genetic architectures across ASD and DD, with significant heterogeneity across these broadly defined NDDs. To explore commonalities in genes that impact such NDDs, we integrated our ASD data with an independent cohort of 91,605 family-based samples encompassing 31,058 offspring with DD, the vast majority of which (>95%) were diagnosed with at least one NDD^11^. Exome sequencing from this DD cohort was recently analyzed by the Deciphering Developmental Disorders (DDD) project using DeNovoWest, a permutation-based frequentist method, and reported exome-wide significant associations between rare SNVs and indels in 252 autosomal genes. However, these analyses did not include case-control samples, rare inherited SNVs and indels, or CNVs. We sought to re-analyze these DDD data to enable direct comparisons between the ASD and DD cohorts using uniform statistical models and significance thresholds between studies.

We applied the extended TADA framework to *de novo* variants in the DD cohort (TADA-DD) and found 309 autosomal genes associated at an FDR threshold of ≤ 0.001, including 94% of the initial 252 autosomal genes in Kaplanis *et al*. 2020^11^ (**Supplementary Table 4.9**, TADA-DD found 477 at FDR ≤ 0.05). We observed the FDR values generated by our implementation of TADA (TADA-DD) to be highly correlated to those derived from the DeNovoWest significance values in Kaplanis *et al*. 2020^11^ (r=0.95, p<10×10-22, **Supplementary Fig. 5**). Given the enrichment of cases with severe and syndromic disorders in this DD cohort compared to the ASD cohort^10,11,61^, the *de novo* PTV, MisB, and MisA counts in this DD cohort showed the expected stronger but overall similar trends in variant enrichment in probands across the top three deciles of LOEUF (**Fig. 5a-b**).

**Fig. 5.**
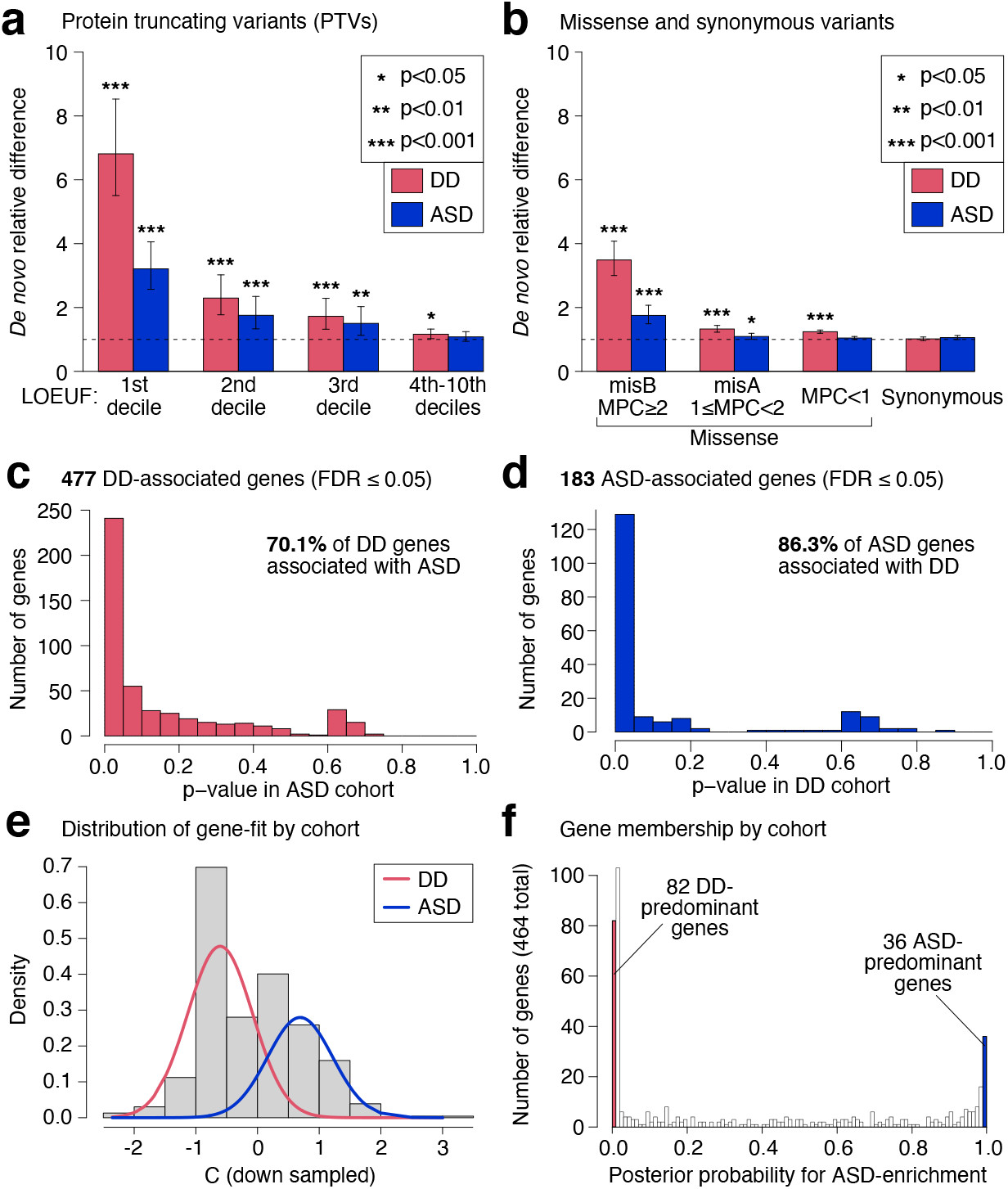
Integration of ASD and DD dataset identifies discovery of general neurodevelopmental disorder genes and heterogeneity. We analyzed 31,058 DD trios from Kaplanis et al. 2020 alongside our assembled ASD cohort to compile a combined dataset of 46,094 NDD cases. a, De novo PTVs are enriched in DD cases, with the effect diminishing as constraint relaxes. The enrichment in DD cases is higher than in the ASD cases, consistent with expectation. b, De novo missense variants are also enriched in DD cases above and beyond the enrichment observed in our ASD cases. c, 477 TADA-DD genes with FDR ≤ 0.05 have non-uniform p-value distributions from TADA-ASD that suggest 70.1% are associated with ASD. d, 183 TADA-ASD genes with FDR ≤ 0.05 have non-uniform p-value distributions from TADA-DD that suggest 86.3% of these genes are associated with DD. e, Using PTV and MisB variant data, we devised a chi-squared statistic, denoted the C statistic, to measure if a gene has more observed variants in one cohort relative to the other. A mixture model was used to deconvolve the commingled distributions. f, We transformed the fitted mixture distribution into posterior probability for ASD enrichment. Using a cutoff of <0.01, we found 82 DD-predominant genes, while using a cutoff of >0.99 we found 36 ASD-predominant genes. Abbreviations: PTV: protein truncating variants; BF: Bayes factor; DD: developmental delay; FDR: false discovery rate. Statistical tests: a-b: binomial test, c-d: limma:: propTrueNull:, e-f: mixture model.

Since the first unbiased whole-genome analyses in ASD^62^, a notable overlap has been observed between genes affecting ASD and those affecting development more broadly, including intellectual development. Thus, it is reasonable to conjecture that integrating evidence for association from ASD and DD cohorts could yield additional genes influencing risk for ASD and broader development. Still, a cardinal rule of such meta-analysis is that the data should not be too heterogeneous. To determine whether the genes identified in the ASD cohort were also associated in the DD cohort, and vice versa, we converted the distribution of TADA q-values to p-values for each study. If the genes identified as associated in the DD cohort were also associated with ASD, the distribution of their ASD association p-values would be skewed toward zero; if they were not associated, the p-values would follow a uniform distribution; and, if some were associated and others were not, the p-values would follow a mixture distribution. Selecting the genes associated in the DD cohort at FDR ≤ 0.05, we evaluated the distribution of their p-values in the ASD cohort. The estimated proportion of associated genes (i.e., mixing parameter) was 0.701 (limma::propTrueNull, **Fig. 5c**), indicating that 70.1% of these genes affect risk for ASD. The converse analysis conditioning on the ASD-associated genes suggests that 86.3% of ASD risk genes have some broad effects on development (limma::propTrueNull, **Fig. 5d**).

Having observed that the results from the ASD and DD cohorts are somewhat complementary, we used the Bayesian framework underpinning TADA to integrate the genetic evidence for each gene across the cohorts by combining the BFs, which is conceptually similar to a frequentist meta-analysis. This combined analysis (TADA-NDD) revealed 373 genes associated with general NDDs at FDR ≤ 0.001 and 662 genes at FDR ≤ 0.05 (**Supplementary Table 4.9**). We observed that 54/373 and 124/662 genes did not meet the matching threshold in either the TADA-ASD or TADA-DD analyses alone, demonstrating a 15-19% increase in yield by combining evidence from these cohorts.

### Heterogeneity of mutation patterns between ASD and DD associated genes

Due to the frequent comorbidity of ASD and DD phenotypes, isolating genes that exert a greater effect on ASD than they do on other DDs has remained challenging. Still, as documented above, 13.7% of the TADA-ASD FDR ≤ 0.05 genes show little evidence for association in the DD cohort at the same threshold (**Fig. 5d**). Moreover, while the remaining 86.3% of the genes are likely pleiotropic^63^, some could have a far greater impact on risk for ASD while others have greater impact on risk for other features of development. To evaluate the overall gene-set heterogeneity between the ASD and DD cohorts, we retained only *de novo* SNVs/indels for independent gene-level BF calculations. For the 373 genes at TADA-NDD FDR ≤ 0.001, we observed a Pearson’s correlation of 0.78 (p=9.1×10^−78^) of the gene-level log BF between the two major subcomponents of the ASD cohort (ASC+SSC versus SPARK), compared to only 0.42 (p=2.4×10^−17^) between the BFs of the ASD and DD cohorts. These observations signal much more consistent gene-level evidence between the ASD subcohorts than between the ASD and DD cohorts (**Supplementary Fig. 6**) and reflect both differential and shared genetic architecture underlying these often-comorbid phenotypes.

Next, we determined if certain genes were observed to be more commonly mutated in one cohort or the other. For these analyses, we first identified 464 “signal genes,” defined as any gene with FDR ≤ 0.05 in either TADA-ASD or TADA-DD from *de novo* PTV and MisB variants (which showed the greatest association and therefore were less likely to be benign). Of these signal genes, 120 belonged to TADA-ASD, 428 to TADA-DD, and 84 to both. Notably, even for the 84 genes significant in both cohorts, we also observed significant variant count heterogeneity (X2=317.6, DF=83, p=3.8×10^−23^) between the cohorts.

Given this heterogeneity in mutational patterns, we sought to capture which of the 464 genes have more variation in either the ASD or the DD cohorts. A common way to assess this is by a standardized chi-squared test statistic (C statistic), but its power to discriminate is abrogated by the much higher burden of risk variants in the DD than ASD samples (**Fig. 5a-b**). We therefore adjusted for the difference in mutational burden between the cohorts by downsampling the DD mutations to be equal to the count of ASD mutations, then computed the C statistic. Genes with fewer than expected mutations in the ASD cohort relative to the DD cohort achieved a negative C statistic; a positive C statistic indicated overrepresentation in the ASD cohort. Next, a mixture model was adopted to disentangle the two commingled distributions, which assigns posterior probabilities that a gene is from the ASD or DD component of the statistical distribution (**Fig. 5e-f, Supplementary Table 4.11**). Using a posterior probability cutoff of greater than 0.99, we find 36 genes to be a part of the ASD mixture component and 82 genes to be a part of the DD component (**Fig. 5f**).

### Mutational enrichment identifies differential neuronal layers impacted by ASD and DD genes

To gain insight into differences in expression of genes identified from the ASD versus DD cohorts, we examined single-cell gene expression patterns from human fetal brains. Two studies provided more than 37,000 cortical cells ranging from 6-27 weeks post-conception^64,65^. To integrate these data, we first removed strong batch effects due to different protocols using cFIT and then obtained integrated factor loadings and gene expression for all measured cells^66^. UMAP plots visualizing all cells from two sources, after removal of batch effects, showed that similar cell types from the different batches group together, while cells unique to either batch were also preserved^66^ (**Fig. 6a, Supplementary Fig. 7**).

**Fig. 6.**
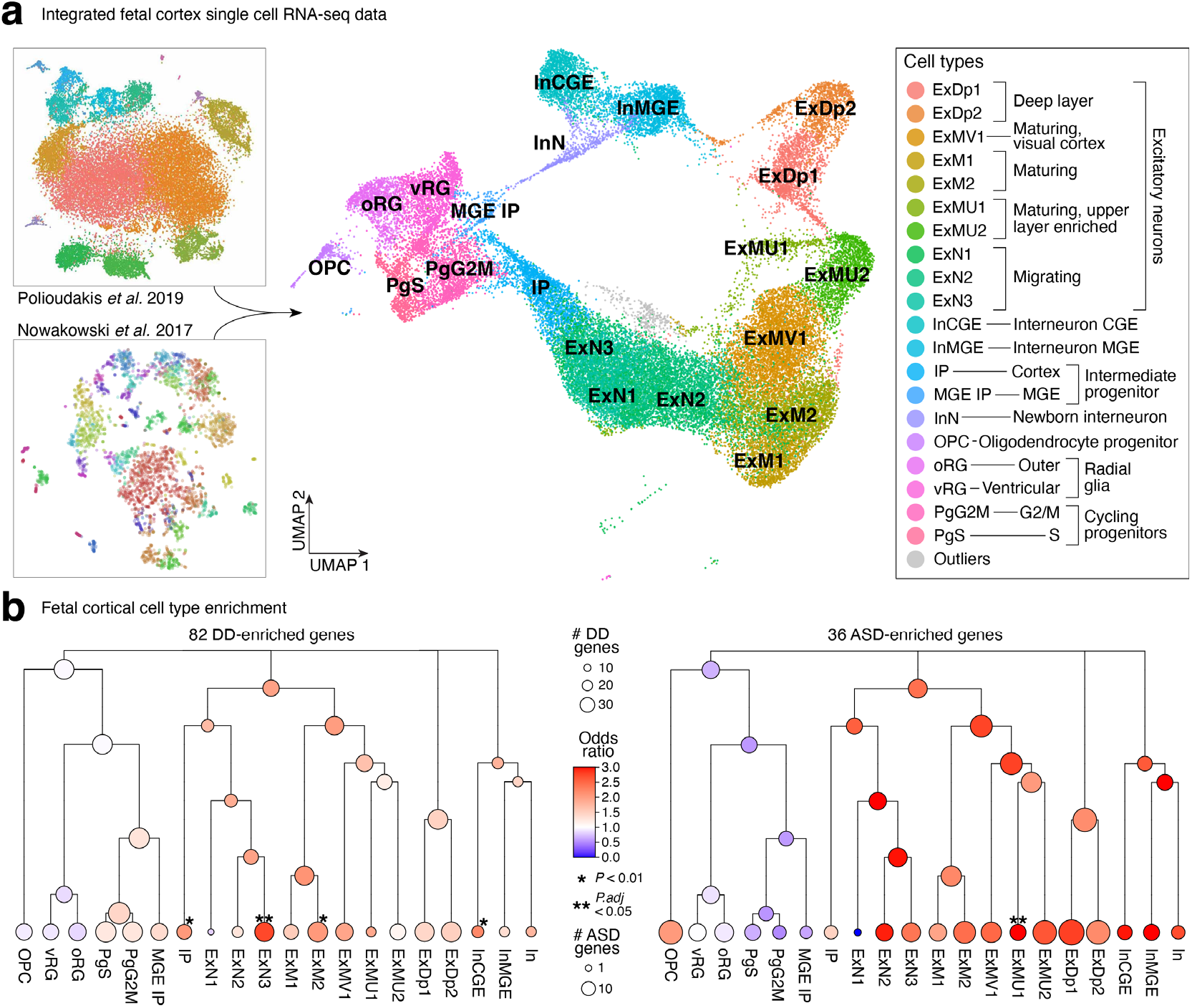
Single-cell data reveals differential neuronal layers impacted by ASD and DD genes. a, A UMAP plot visualization after integrating two studies (Nowakowski et al. 2017; Polioudakis et al. 2019) that provided single-cell gene expression of human fetal brains consisting of 37,000 cortical cells 6-27 weeks post-conception. Similar cell types from the two batches group together while preserving cells unique to either study. b, Both ASD- and DD-predominant genes (right and left, respectively) were found to be enriched in interneurons and excitatory neurons compared to glial cells. Compared to DD-predominant genes, ASD-predominant genes are relatively more neuron-enriched than progenitor-enriched. Abbreviations: UMAP: Uniform Manifold Approximation and Projection Statistical tests: b: Fisher’s exact

With the integrated data, we applied unsupervised clustering to identify cell subtypes in the context of a hierarchical tree to illustrate the relationships of major and minor cell type clusters. Using the MRtree method^67^, we observed that cells of each labeled type were merged across both datasets into common clusters. Visualizing the tree, the major branches corresponded to glial and progenitor cells, excitatory neurons, deep layer enriched excitatory neurons, and inhibitory neurons. Likewise, minor splits reflected the expected relationships between cell types (**Supplementary Fig. 8a**). Based on the trajectory analysis of Polioudakis *et al*.^65^, the ExN clade is less mature than the ExM clade, which in turn is less mature than the ExMU clade.

We subsequently assessed any differentials in strength of enrichment of the DD- and ASD-predominant risk genes within cell clusters for genes meeting the posterior probability 0.99 threshold (**Fig. 5f**). Of the 36 genes classified as ASD-predominant, 18 were expressed in these cell types; and, of the 82 classified as DD-predominant genes, 51 were expressed. Using odds ratio to reflect the strength of signal, we observed that both DD- and ASD-predominant genes were enriched in both interneurons and excitatory neurons compared to glial and progenitor cells (**Fig. 6b, Supplementary Fig. 9, Supplementary Tables 4.12-4.13**). While the enrichment was broadly distributed, the signal for DD-predominant enrichment tended to be found for cell types appearing early in the lineages of neurons. DD-predominant genes highlight four cell types, namely intermediate progenitors (IP), precursors of interneurons from the caudal ganglionic eminence (InCGE), migrating excitatory neurons (ExN3), and maturing excitatory neurons (ExM2). By contrast, ASD-predominant genes are strongly enriched in only one cell type, maturing excitatory neurons (ExMU1) and its clade. In keeping with a more mature neuron type, the ASD-predominant genes play roles in cytoskeleton organization, synaptic signaling, and activity-dependent programs. Our results regarding ASD agree with the conclusion of Polioudakis *et al*.^65^, which was based on a subset of these data. If we judge enrichment solely by significance after Bonferroni correction for 21 cell types, these conclusions still hold: ExMU1 remained significant for enrichment of ASD-predominant genes; likewise ExN3 remained significant for enrichment of DD-predominant genes. Our results are consistent with DD-predominant genes being expressed earlier in development and in less mature cells than ASD-predominant genes.

### Emergence of shared ASD schizophrenia risk genes

The genetic risk for ASD has also been suggested to overlap with that of other neuropsychiatric disorders, especially schizophrenia^68^, such that the joint study of ASD and schizophrenia might reveal additional insights into both disorders. The Schizophrenia Exome Meta-Analysis (SCHEMA) Consortium recently analyzed exome sequencing from 24,248 schizophrenia cases and 97,322 controls, identifying 10 genes where ultra-rare coding variants were associated with schizophrenia after Bonferroni correction, and 244 genes at p < 0.01^13^. We compared our ASD- and NDD-associated genes to the SCHEMA findings to determine if there was any overlap between ASD and schizophrenia at the level of individual risk genes and, if so, whether it was related to ASD-DD overlap. Among the 71 ASD genes we discovered at an FDR ≤ 0.001, we found that 63 show an association with DD (using FDR ≤ 0.05, based on TADA-DD), and 8 show an association with schizophrenia (using p ≤ 0.01, based on values reported by Singh *et al*.). If the two associations were independent, we would expect ∼7 of the 8 ASD-schizophrenia genes to also show an association with DD (based on 63/71 = ∼88% of the ASD genes overlapping DD). However, we find 4 of the ASD-schizophrenia genes (*NRXN1, ANK2, BRSK2*, and *DSCAM*) lack an association with DD, which is a significant overrepresentation compared to random chance alone (p = 0.008, binomial test). This suggests that one subset of ASD risk genes may overlap DD, while a different subset overlaps schizophrenia.

We also analyzed these findings with an alternative approach: the distribution of genes in **Fig. 5e** (the 464 genes with FDR ≤ 0.05 for ASD and/or DD in our heterogeneity analysis) gives 36 ASD-enriched genes and 82 DD-enriched genes. Of the 244 genes identified by SCHEMA as schizophrenia-associated at p ≤ 0.01, 6 genes (*ANK2, ASH1L, BRSK2, CGREF1, DSCAM*, and *NRXN1*) overlap the 36 ASD genes, while only 3 (*ATP2B1, GRIN2A*, and *HIST1H1E*) overlap the 82 DD genes. If we compare to the null hypothesis that each of the genes in our TADA model has an equal chance of being schizophrenia-associated, then the ASD-schizophrenia overlap is significantly enriched (p = 8.5×10^−6^, binomial test), while the DD-schizophrenia overlap is not (p = 0.10, binomial test). The two outcomes (6/36 vs. 3/82) are also significantly different when compared to each other (p = 0.023, Fisher’s exact test), reinforcing the idea that the shared genetic risk between ASD and schizophrenia may be distinct to that observed between ASD and DD.

## Discussion

We report here a gene discovery effort in ASD that integrates coding SNVs, indels, and exome-based CNVs in ASD. These analyses revealed an allelic spectrum associated with ASD that was dominated by *de novo* PTVs, damaging missense variants, and deletions within genes that display evidence of intolerance to loss-of-function variation in the general population. Nonetheless, of the 71 genes identified as associated at FDR ≤ 0.001, all included association evidence from multiple inheritance classes and 10 displayed the strongest evidence from *de novo* missense variants and duplications, including one gene (*PLXNA1*) with domain-specific transmission distortion. The proportion of log BF evidence arising from PTVs and DELs steadily decreased from 69.2% among the 71 TADA-ASD risk genes to 51.1% and 41.0% for FDR ranges of (0.001, 0.01) and (0.01, 0.05) respectively, while inherited evidence increased from 4.5% to 10.1% and to 20% for the same FDR ranges, indicating that future discoveries are likely to arise from the more subtle effects associated with missense and inherited variation as sample sizes continue to increase.

This study also presents the largest exploration to date of individual gene and multi-exon resolution CNVs from exome sequencing and their contributions to ASD architecture. In a subset of the SSC quartets in this study, we demonstrated that >80% of all CNVs discovered by WGS and spanning three or more exons could be recaptured by exome-based CNV detection, while >90% of all predicted *de novo* CNVs from exome sequencing at this resolution were confirmed by WGS and/or molecular validation. We readily recapitulated the observed excess of CNVs in known GD regions characterized from microarray studies, and quantified the relative effect size of CNVs outside of GD regions in comparison to PTVs. Prior studies have reported an enrichment of CNVs of maternal origin in ASD probands^57^. Here, we found a strong bias in the gamete-of-origin giving rise to de novo CNVs based on the mechanism of formation. Most NAHR-mediated CNVs across all GDs arose on the maternal chromosome (e.g., 95% of 16p11.2 CNVs), whereas the majority of all CNVs, which are driven by non-homologous or microhomology mediated repair, were preferentially paternal in origin and consistent with WGS analysis in controls^48^. We further highlighted the relative value of gene-level CNV analyses, as disruption of one or more highly constrained genes conferred comparable risk to alteration of dosage-sensitive GD loci. Overall, approximately 8% of the BF association evidence in ASD risk genes was derived from CNVs, while 0.95% of all cases and 0.02% of unaffected siblings harbored a *de novo* CNV that altered one of the 373 broadly defined NDD risk genes. These results emphasize the value of routine joint analysis of all classes of genomic variation in gene discovery analyses, and the relative impact of gene level CNV analyses in diagnostic testing.

By analyzing both the DD and ASD datasets under the same framework, we were also able to directly quantify the overlapping and distinct genetic architectures across these datasets. We showed the increased rate of *de novo* PTVs and damaging missense variants in the DD cohort in both raw variant enrichments as well as in association statistics. Applying the same statistical model to both the DD and ASD datasets independently reinforced that the analytic framework and statistical thresholds used were robust and highly correlated with the permutation-based approach applied in the DDD study. Integrating the two cohorts together with TADA, we implicated 373 genes associated with risk across a spectrum of syndromic and non-syndromic NDDs at FDR ≤ 0.001 (662 risk genes at FDR ≤ 0.05). Among these 373 genes, 54 genes were unique to the joint analyses and were not captured by either dataset alone.

We expect these gene sets to shed light on the neurobiological origins of ASD, including its key features, namely deficits in social interaction and the presence of repetitive behaviors and/or interests, versus broader DD phenotypes. One might reasonably ask that if there is substantial overlap between the genes implicated in NDDs writ large and those implicated directly in ASD, why is it important to focus on ASD at all? To see how a larger set of risk genes and one originating from analysis of an ASD cohort could be important, consider two of the 183 genes, specifically *ARID1B* and *DSCAM*. While both are highly associated with ASD, with FDRs of 5.5×10^−12^ and 3.7×10^−4^ respectively, statistical evidence for *ARID1B*’s association with ASD is stronger. Nine of 15,036 ASD probands carry PTV or misB *de novo* mutations in *ARID1B*, while 4 of 15,036 carry such mutations in *DSCAM*. These genes, however, are also distinguished by their association with broader phenotypes. Individuals with mutations in *ARID1B* are often given a diagnosis of Coffin-Siris syndrome and, according to recent work^69^, they typically have “ID, feeding difficulties, laryngomalacia, speech delay, motor delay, hypertrichosis, and cryptorchidism.” While some individuals with mutations in *ARID1B* also have comorbid ASD, it is only one of a wide range of rarer phenotypes^69^. *ARID1B*’s profound impact on development is apparent by the contrast of *de novo* mutations in the DD and ASD cohorts: 132 carriers out of 31,058 DD probands versus 9 carriers out of 15,036 ASD probands, a sevenfold higher rate in DD. This raises a challenge for neurobiologists. If they perturb *ARID1B* and identify neurodevelopmental features associated with that perturbation, are those features relevant to ASD or are they non-specific features of DD? One might instead select *DSCAM* because its evidence for association arose solely from the ASD cohort; no PTV or misB mutations were reported from 31,058 DD probands. Yet, as we develop here, there is evidence *DSCAM* is also involved in risk for schizophrenia, so it is not uniquely an “autism gene” and studies such as ours continue to demonstrate the pleiotropic consequences of many genes implicated in ASD and NDD risk. To identify the key neurobiological features of ASD will likely require convergence of evidence from many ASD genes and studies. Careful selection of candidates among the genes implicated here based on their mutational and functional features could inform these future studies.

We have taken a step in that direction with this study. First, we asked which of the ASD and DD genes had a predominance of mutations in the ASD versus DD cohorts. Gene lists identified from cohorts ascertained for ASD and DD show substantial overlap, with roughly 70 to 90% of genes contributing risk for both disorders. This genetic result matches clinical experience with individuals who carry damaging DNA variants in conserved, developmentally important genes. This population tends to show substantial comorbidity between the two diagnoses, as well as phenotypic data, with higher rates of cognitive impairment and walking delay in individuals with ASD^10,11,15,22^. We would predict elevated rates of social impairment in those with DD who carry ASD risk-mediating variants, although this remains to be assessed. Such an assessment is important because, while overlap between genes identified in ASD and DD is substantial, the risk mediated to each disorder is not equal for many of the genes. Broadly, genes expressed at earlier stages of cortical development, such as progenitor genes, display greater DD enrichment, while those expressed later, such as maturing neurons, lean towards ASD. This is consistent with the expectation that earlier and more generalized impairment leads to severe global developmental delay, and later, neuron-specific impairments affect more isolated developmental domains such as social communication. This is supported by our analyses incorporating single-cell genomic data, where we identify ExMu1 among maturing excitatory neurons as preferentially perturbed among ASD-predominant risk genes, while highlighting ExN3, a migrating excitatory neuron, in DD-predominant risk genes.

In conclusion, we have greatly expanded the rare variant information that is simultaneously accounted for in our statistical framework, including the use of accurate WES-based CNVs, to significantly increase the number of genes we can implicate for ASD and NDD risk. We have also used our framework to study the shared and distinct genetic risks between ASD and related NDDs, highlighting differential enrichment of associated genes at different neuronal timepoints. Importantly, much of the work presented in this study was powered by the large-scale collection of ASD datasets from the SPARK, ASC, SSC, and NHGRI Centers for Common Disease Genetics, and the commitment of these programs to make these data rapidly accessible to all qualified individuals. These studies have catalyzed a rapid evolution in genetic architecture studies in ASD, including a number of recent preprints that have leveraged these data for analyses of de novo, ultra-rare, and rare inherited variants in ASD, the combined impact of rare and common variant polygenic risk across males and females, and combined gene discovery from ASD and DD datasets^15,70–72^. As sample sizes continue to grow rapidly, we expect that the framework presented here will continue to yield returns in both gene discovery and improved understanding of the differential risks to the disorders on the neurodevelopmental and neuropsychiatric spectrum posed by variants within the same genes.

## Data Availability

All data used in this study are deposited at:
Repository/DataBank Accession: NHGRI AnVIL
Accession ID: phs000298
Databank URL: https://anvilproject.org/data
Repository/DataBank Accession: Simons Foundation for Autism Research Initiative SFARIbase
Accession ID: SPARK/Regeneron/SPARK_WES_2/
Databank URL: https://www.sfari.org/resource/spark/

## ACKNOWLEDGEMENTS

We thank all of the individuals who participated in this research. We also thank all contributing investigators to the consortia datasets used here from the Autism Sequencing Consortium (ASC), the Simons Simplex Collection (SSC), Simons Powering Autism Research for Knowledge (SPARK) project, the iPSYCH project, the Deciphering Developmental Disorders (DDD) project, and Schizophrenia Exome Meta-Analysis (SCHEMA).

This work was supported by grants from the Simons Foundation for Autism Research Initiative (SSC-ASC Genomics Consortium #574598 to S.J.S., #575097 to B.D. and K.R., #573206 to M.E.T. and M.J.D., #571009 to J.D.; the SPARK project and SPARK analysis projects #606362 and #608540 to M.E.T., M.J.D., J.B., B.D., K.R., S.J.S.; SFARI #402281 to S.J.S., M.W.S., B.D., and K.R. and #647371 to S.J.S.), NHGRI (HG008895 to M.J.D., S.G., M.E.T.), NIMH (MH115957 to M.E.T., MH111658 and MH057881 to B.D., MH111661 and MH100233 to J.D.B., R01 MH109900 to K.R., MH111660 to M.J.D., and MH111662 and MH100027 to S.J.S. and M.W.S.), NICHD (HD081256 to M.E.T.), AMED (JP21WM0425007 to N.O). and the Seaver Foundation. J.M.F. was supported by an Autism Speaks Postdoctoral Fellowship and R.L.C. was supported by NSF GRFP #2017240332.

## AUTHOR CONTRIBUTIONS

**Study design**: M.E.T, S.J.S, K.R., B.D., M.J.D, J.D.B, S.B.G; **Sample contribution and data generation**: M.E.T., M.J.D., J.D.B., S.D.R., S.B.G., S.D., C.C., C.R.S., M.B, A.B, B.H.Y.C., M.L.C., E.D, G.B.F., J.J.G., G.E.H., I.H., P.M., D.S.M., M.R.P., A.M.P., A.R., F.T., E.T., G.C., M.C.Y.C., C.F., E.G., A.C.G., E.H., S.L.L., C.L., Y.L., R.N., L.P., M.P-V., I.P., E.R., R.S., M.S., C.I.C.S., S.T., J.Y.T.W., M.H.C.Y.; **Project management of coordination**: M.E.T., S.J.S., M.J.D., J.D.B., S.D.R., L.S., B.M., C.R.S., C.S., B.C., J.J.G.; **Methods development and analysis**: M.E.T., S.J.S., K.R., B.D., M.J.D., D.J.C., A.S., A.L., L.L., S.K.L., L.G., E.B., M.B., L.K., S.Dong, R.L.C., H.B., M.P., F.K.S., J.M.F.; **Writing**: M.E.T., S.J.S., K.R., B.D., M.J.D., J.D.B., H.B., M.P., K.F.S., J.M.F.

