## Supplement for "Rare coding variation illuminates the allelic architecture, risk genes, cellular expression patterns, and phenotypic context of autism"

### Autism Sequencing Consortium (ASC)

Branko Aleksic 44, Joon-Yong Anney 82, Richard Anney 83, Mykyta Artomov 1,2,4,64, Mafalda Barbosa 14,17, Elisa Benetti 31,32, Catalina Betancur 84, Monica Biscaldi-Schafer 54, Somer Bishop 9, Anders D. Børglum 45,46,47,48, Harrison Brand 1,2,3,7, Michael S. Breen 12,13,14, Alfredo Brusco 18,19, Joseph D. Buxbaum 12,14,40, Gabriele Campos 29, Angel Carracedo 52,65, Marcus C.Y. Chan 20, Andreas G. Chiocchetti 54, Brian H.Y. Chung 20, A. Ercument A. Cicek 43,85, Rachel Cohen 12,13, Brett Collins 12,13,14, Ryan L. Collins 1,2,3,8, Edwin H. Cook 49, Hilary Coon 50,51, Michael L. Cuccaro 21, David J. Cutler 38, Bernardo Dalla Bernardina 86, Mark J. Daly 1,2,4,5,41,42, Silvia De Rubeis 12,13,14,39, Bernie Devlin 10, Caroline Dias 66,87, Ryan N. Doan 66, Enrico Domenici 22, Shan Dong 9, Chiara Fallerini 31,32, Montserrat Fernández-Prieto 52,53, Giovanni Battista Ferrero 23, Christine M. Freitag 54, Menachem Fromer 2, Jack M. Fu 1,2,3, Louise Gallagher 88, Jay J. Gargus 24, Sherif Gerges 1,2,4,64, Daniel Geschwind 72,89, Michael Gill 88, Michael Gilson 9, Elisa Giorgio 18, Ana Cristina Girard 29, Javier González-Peñas 60, Jakob Grove 45,46,47,48, Elizabeth E. Guerrero 26, Stephen Guter 49, Danielle Halpern 12,13, Emily Hansen-Kiss 25, Xin He 90, Gail E. Herman 25, Irva Hertz-Picciotto 26, David M. Hougaard 45,55, Christina M. Hultman 16, Iuliana Ionita-Laza 91, Suma Jacob 49, Jesslyn Jamison 12,13, Magdalena Janecka 12,13,92, Astanand Jugessur 76, Miia Kaartinen 61, Lambertus Klei 10, Gun Peggy Knudsen 76, Alexander Kolevzon 12,13,56, Itaru Kushima 44,67, So Lun Lee 20, Terho Lehtimäki 68, Tess Levy 12,13, Yue Li 6,43, Lindsay Liang 9, Elaine T. Lim 64, Carla Lintas 36, W. Ian Lipkin 91, Alicia Ljungdahl 9, Caterina Lo Rizzo 32, Fátima Lopes 27, Yunin Ludena 26, Patricia Maciel 27, Per Magnus 76, Behrang Mahjani 12,13,16, Nell Maltman 49, Marianna Manara 32, Dara S. Manoach 28, Gal Meiri 69,70, Idan Menashe 57,58, Judith Miller 71,72, Nancy Minshew 10, Eric M. Morrow 93, Matthew Mosconi 73, Pierandrea Muglia 94, Benjamin M. Neale 2,4,5, Rachel Nguyen 24, Utku Norman 85, Norio Ozaki 44,74, Aarno Palotie 2,5,42,59, Mara Parellada 60, Maria Rita Passos-Bueno 29, Lisa Pavinato 18, Minshi Peng 6, Margaret Pericak-Vance 21, Antonio M. Persico 30, Isaac Pessah 26,37, Kaija Puura 61, Abraham Reichenberg 12,13,14,75, Alessandra Renieri 31,32,33, Evelise Riberi 23, Elise B. Robinson 2,4,5, Kathryn Roeder 6,43, Kaitlin E. Samocha 1,2, Stephan J. Sanders 9, Sven Sandin 12,13,16, Susan L. Santangelo 34,95, F. Kyle Satterstrom 2,4,5, Gerry Schellenberg 96, Stephen W. Scherer 62,63, Sabine Schlitt 54, Rebecca Schmidt 26, Lauren Schmitt 49, Katja Schneider-Momm 54, Tarjinder Singh 2,4,5,64,97, Paige M. Siper 12,13,14, Laura Sloofman 12,13,14, Moyra Smith 24, Gabriela Soares 98, Claudia I.C. Souza 29, Matthew W. State 9, Christine R. Stevens 2,4,5, Camilla Stoltenberg 76, Pål Suren 76, Ezra Susser 99,100, James S. Sutcliffe 101,102, John A. Sweeney 77, Peter Szatmari 103, Michael E. Talkowski 1,2,3,4,8, Flora Tassone 26,34, Karoline Teufel 54, Audrey Thurm 104, Elisabetta Trabetti 35, Slavica Trajkova 18, Maria del Pilar Trelles 12,13, Christopher A. Walsh 80,105,106, Brie Wamsley 72, Jaqueline Y.T. Wang 29, Yuting Wei 6,43, Lauren A. Weiss 9, Donna Werling 107, Emilie M. Wigdor 2,4,5, Jeremy A. Willsey 9,108,109, Mullin H.C. Yu 20, Timothy W. Yu 110, Ryan Yuen 62, Michael E. Zwick 38

### iPSYCH-BROAD Consortium

Esben Agerbo 45,111,112, Thomas Damm Als 45,113,114, Vivek Appadurai 45, Marie Bækved-Hansen 45,115, Rich Belliveau 4, Carsten Bøcker Pedersen 45,111,112, Anders D. Børglum 45,46,47,48, Alfonso Buil 45,116, Jonas Bybjerg-Grauholm 45,55, Caitlin E. Carey 2,4,5, Felecia Cerrato 4, Kimberly Chambert 4, Claire Churchhouse 2,4,5, Søren Dalsgaard 45,113,114, Mark J. Daly 1,2,4,5,41,42, Ditte Demontis 45,113, Ashley Dumont 4, Mads Engel Hauberg 45,113,114, Marianne Giørtz Pedersen 45,111,112, Jacqueline Goldstein 2,4,5, Jakob Grove 45,46,47,48, Christine S. Hansen 45,55,116, Mads V. Hollegaard 55, David M. Hougaard 45,55, Daniel P. Howrigan 4,5, Hailiang Huang 4,5, Julian Maller 4,117, Alicia R. Martin 2,4,5, Joanna Martin 4,16,118, Manuel Mattheisen 45,113,119,120, Jennifer Moran 4, Ole Mors 45,121, Preben Bo Mortensen 45,46,111,112, Benjamin M. Neale 2,4,5, Merete Nordentoft 45,122, Jonatan Pallesen 45,113,114, Duncan S. Palmer 4,5, Timothy Poterba 2,4,5, Jesper Buchhave Poulsen 45,55, Stephan Ripke 2,5,123, Elise B. Robinson 2,4,5, F. Kyle Satterstrom 2,4,5, Andrew J. Schork 116, Christine R. Stevens 2,4,5, Wesley K. Thompson 116,124, Patrick Turley 4,5, Raymond K. Walters 4,5, Thomas Werge 45,116,125, Emilie M. Wigdor 2,4,5

### Broad Institute Center for Common Disease Genomics (Broad-CCDG)

Felecia Cerrato 4, Claire Churchhouse 2,4,5, Caroline Cusick 4, Mark J. Daly 1,2,4,5,41,42, Anne Feng 78,79, Stacey B. Gabriel 15, Namrata Gupta 2, Sekar Kathiresan 1,80,81, Amit Khera 1, Eric Lander 80, Robert Maier 2,4,5, Benjamin M. Neale 2,4,5, Candace Patterson 2, Christine R. Stevens 2,4,5, Michael E. Talkowski 1,2,3,4,8

### Affiliations

1. Center for Genomic Medicine, Massachusetts General Hospital, Boston, MA 02114, USA; 2. Program in Medical and Population Genetics, Broad Institute of MIT and Harvard, Cambridge, MA 02142, USA; 3. Department of Neurology, Massachusetts General Hospital and Harvard Medical School, Boston, MA 02114, USA; 4. Stanley Center for Psychiatric Research, Broad Institute of MIT and Harvard, Cambridge, MA 02142, USA; 5. Analytic and Translational Genetics Unit, Department of Medicine, Massachusetts General Hospital, Boston, MA 02114, USA; 6. Department of Statistics and Data Science, Carnegie Mellon University, Pittsburgh, PA 15213, USA; 7. Pediatric Surgical Research Laboratories, Massachusetts General Hospital, Boston, MA 02114, USA; 8. Program in Bioinformatics and Integrative Genomics, Harvard Medical School, Boston, MA 02115, USA; 9. Department of Psychiatry, UCSF Weill Institute for Neurosciences, University of California San Francisco, San Francisco, CA 94143, USA; 10. Department of Psychiatry, University of Pittsburgh School of Medicine, Pittsburgh, PA 15213, USA; 11. Data Sciences Platform, The Broad Institute of MIT and Harvard,Cambridge, MA 02142, USA; 12. Seaver Autism Center for Research and Treatment, Icahn School of Medicine at Mount Sinai, New York, NY 10029, USA; 13. Department of Psychiatry, Icahn School of Medicine at Mount Sinai, New York, NY 10029, USA; 14. The Mindich Child Health and Development Institute, Icahn School of Medicine at Mount Sinai, New York, NY 10029, USA; 15. Genomics Platform, The Broad Institute of MIT and Harvard, Cambridge, MA 02142, USA; 16. Department of Medical Epidemiology and Biostatistics, Karolinska Institutet, 171 77 Stockholm, Sweden; 17. Department of Genetics and Genomic Sciences, Icahn School of Medicine at Mount Sinai, New York, NY 10029, USA; 18. Department of Medical Sciences, University of Torino, Turin, 10126, Italy; 19. Medical Genetics Unit, "Città della Salute e della Scienza" University Hospital, Turin, 10126, Italy; 20. Department of Pediatrics & Adolescent Medicine, Duchess of Kent Children's Hospital, The University of Hong Kong, Hong Kong Special Administrative Region 999077, China; 21. The John P Hussman Institute for Human Genomics, The University of Miami Miller School of Medicine, Miami, FL 33136, USA; 22. Department of Cellular, Computational and Integrative Biology, University of Trento, 38122 Trento, Italy; 23. Department of Public Health and Pediatrics, University of Torino, Turin, 10126, Italy; 24. Center for Autism Research and Translation, University of California Irvine, Irvine, CA 92697, USA; 25. The Research Institute at Nationwide Children's Hospital, Columbus, OH 43205, USA; 26. MIND (Medical Investigation of Neurodevelopmental Disorders) Institute, University of California Davis, Davis, CA 95616, USA; 27. Life and Health Sciences Research Institute, School of Medicine, University of Minho, Campus de Gualtar, 4710-057 Braga, Portugal; 28. Department of Psychiatry, Massachusetts General Hospital and Harvard Medical School, Boston, MA 02114, USA; 29. Centro de Pesquisas sobre o Genoma Humano e Células tronco, Instituto de Biociências, Universidade de São Paulo, São Paulo, 05508090, Brazil; 30. Interdepartmental Program "Autism 0-90", "Gaetano Martino" University Hospital, University of Messina, Messina I-98125, Italy; 31. Med Biotech Hub and Competence Center, Department of Medical Biotechnologies, University of Siena, Ital; 32. Medical Genetics, University of Siena, 53100 Siena, Italy; 33. Genetica Medica, Azienda Ospedaliera Universitaria Senese, 53100 Siena, Italy; 34. Department of Biochemistry and Molecular Medicine, University of California Davis, School of Medicine, Sacramento, CA 95817, USA; 35. Department of Neurosciences, Biomedicine and Movement Sciences, Section of Biology and Genetics, University of Verona, 37134 Verona, Italy; 36. Service for Neurodevelopmental Disorders, University Campus Bio-medico of Rome, 00128 Rome, Italy; 37. Department of Molecular Biosciences, University of California Davis, School of Veterinary Medicine, Davis, CA 95616, USA; 38. Department of Human Genetics, Emory University School of Medicine, Atlanta, GA 30322 USA; 39. Friedman Brain Institute, Icahn School of Medicine at Mount Sinai, New York, NY 10029, USA; 40. Department of Neuroscience, Icahn School of Medicine at Mount Sinai, New York, NY 10029, USA; 41. Department of Genetics, Harvard Medical School, Boston, MA 02115, USA; 42. Institute for Molecular Medicine Finland (FIMM), University of Helsinki, 00290 Helsinki, Finland.; 43. Computational Biology Department, Carnegie Mellon University, Pittsburgh, PA 15213, USA; 44. Department of Psychiatry, Graduate School of Medicine, Nagoya University, Nagoya 466-8550, Japan; 45. The Lundbeck Foundation Initiative for Integrative Psychiatric Research, iPSYCH, 8000 Aarhus, Denmark; 46. Center for Genomics and Personalized Medicine, 8000 Aarhus, Denmark; 47. Department of Biomedicine - Human Genetics, Aarhus University, 8000 Aarhus, Denmark; 48. Bioinformatics Research Centre, Aarhus University, 8000 Aarhus, Denmark; 49. Institute for Juvenile Research, Department of Psychiatry, University of Illinois at Chicago, Chicago, IL 60608, USA; 50. Department of Internal Medicine, University of Utah, Salt Lake City, UT 84132, USA; 51. Department of Psychiatry, Huntsman Mental Health Institute, University of Utah, Salt Lake City, UT 84108, USA; 52. Grupo de Medicina Xenómica, Centro de Investigación en Red de Enfermedades Raras (CIBERER), CIMUS, Universidade de Santiago de Compostela, 15782 Santiago de Compostela, Spain; 53. Neurogenetics group, Instituto de Investigación Sanitaria de Santiago (IDIS-SERGAS), 15706 Santiago de Compostela, Spain; 54. Department of Child and Adolescent Psychiatry, Psychosomatics and Psychotherapy, Goethe University Frankfurt, 60528 Frankfurt, Germany; 55. Center for Neonatal Screening, Department for Congenital Disorders, Statens Serum Institut, 2300 Copenhagen, Denmark; 56. Department of Pediatrics, Icahn School of Medicine at Mount Sinai, New York, NY 10029, USA; 57. Department of Public Health, Ben-Gurion University of the Negev, Beer-Sheva, 841050, Israel; 58. National Autism Research Center of Israel, Ben-Gurion University of the Negev, 8410501, Beer-Sheva, Israel; 59. Psychiatric & Neurodevelopmental Genetics Unit, Department of Psychiatry, Massachusetts General Hospital, Boston, MA 02114, USA; 60. Department of Child and Adolescent Psychiatry, Hospital General Universitario Gregorio Marañón, IiSGM, CIBERSAM, School of Medicine Complutense University, 28009 Madrid, Spain; 61. Department of Child Psychiatry, Tampere University and Tampere University Hospital, 33520 Tampere, Finland; 62.

Program in Genetics and Genome Biology, The Centre for Applied Genomics, The Hospital for Sick Children, Toronto, ON M5G 0A4, Canada; 63. Department of Molecular Genetics and McLaughlin Centre, University of Toronto, Toronto, ON M5S, Canada; 64. Harvard Medical School, Boston, MA 02115, USA; 65. Fundación Pública Galega de Medicina Xenómica, Servicio Galego de Saúde (SERGAS), 15706 Santiago de Compostela, Spain; 66. Division of Genetics and Genomics, Boston Children's Hospital, Boston, MA 02115, USA; 67. Medical Genomics Center, Nagoya University Hospital, Nagoya 466-8550, Japan; 68. Department of Clinical Chemistry, Fimlab Laboratories and Finnish Cardiovascular Research Center-Tampere, Faculty of Medicine and Health Technology, Tampere University, 33520 Tampere, Finland; 69. The Azrieli National Center for Autism and Neurodevelopment Research, Ben-Gurion University of the Negev, 8410501, Beer-Sheva, Israel; 70. Pre-School Psychiatry Unit, Soroka University Medical Center, 8457108, Beer Sheva, Israel; 71. Children's Hospital of Philadelphia, Phladelphia, PA 19104; 72. Department of Psychiatry, University of Utah, Salt Lake City, UT 84108, USA; 73. Life Span Institute and Kansas Center for Autism Research and Training, University of Kansas, Lawrence, KS 66045; 74. Institute for Glyco-core Research (iGCORE), Nagoya University, Furo-cho, Chikusa-ku, Nagoya 464-8601, Japan; 75. Department of Environmental Medicine and Public Health, Icahn School of Medicine at Mount Sinai, New York, NY 10029, USA; 76. Norwegian Institute of Public Health, Oslo, 0213, Norway; 77. Department of Psychiatry, University of Cincinnati, Cincinnati OH 45219 USA; 78. Department of Epidemiology, Harvard T.H. Chan School of Public Health, 655 Huntington Avenue, Building II 2nd Floor, Boston, MA 02115, USA; 79. Metabolomics Platform,Broad Institute of Harvard and MIT, 415 Main St, Cambridge, MA 02142, USA; 80. The Broad Institute of MIT and Harvard, Cambridge, MA 02142, USA; 81. Verve Therapeutics, Cambridge, MA 02139, USA; 82. School of Biosystem and Biomedical Science, College of Health Science, Korea University, Seoul 02841, Republic of Korea; 83. Division of Psychological Medicine & Clinical Neurosciences, MRC Centre for Neuropsychiatric Genetics & Genomics, Cardiff University, Cardiff CF24 4HQ, United Kingdom; 84. Sorbonne Université, INSERM, CNRS, Neuroscience Paris Seine, Institut de Biologie Paris Seine, 75005 Paris, France; 85. Computer Engineering Department, Bilkent University, 06800 Ankara, Turkey; 86. Child Neuropsychiatry Department, Department of Surgical Sciences, Dentistry, Gynecology and Pediatrics. University of Verona, 37134 Verona, Italy; 87. Department of Developmental Medicine, Boston Children's Hospital, Boston, MA 02115, USA; 88. Department of Psychiatry, School of Medicine, Trinity College Dublin, Dublin, D02 VR66, Ireland; 89. Center for Autism Research and Treatment and Program in Neurobehavioral Genetics, Semel Institute, David Geffen School of Medicine, University of California Los Angeles, Los Angeles, CA 90024, USA; 90. Department of Human Genetics, University of Chicago, Chicago, IL 60637, USA; 91. Department of Biostatistics, Columbia University, New York, NY 10032, USA; 92. The Mindich Child Health and Development Institute, Icahn School of Medicine at Mount Sinai, New York, NY 10029, USA.; 93. Department of Molecular Biology, Cell Biology and Biochemistry and Department of Psychiatry and Human Behavior, Brown University, Providence, RI 02912, USA; 94. UCB Pharma, 1420 braine-l'alleud, Belgium; 95. Maine Medical Center and Maine Medical Center Research Institute, Portland, ME 04074, USA; 96. Department of Pathology and Laboratory Medicine, University of Pennsylvania School of Medicine, Philadelphia, PA 19104, USA; 97. Center for Genomic Medicine, Department of Medicine, Massachusetts General Hospital, Boston, MA 02114, USA; 98. Center for Medical Genetics Dr. Jacinto Magalhães, National Health Institute Dr. Ricardo Jorge, 4000-053 Porto, Portugal; 99. New York State Psychiatric Institute, New York, NY 10032, USA; 100. Department of Epidemiology, Mailman School of Public Health, Columbia University, Epidemiology, Mailman School of Public Health, Columbia University, New York, NY 10032, USA; 101. Department of Molecular Physiology & Biophsysics and Psychiatry, Vanderbilt University School of Medicine, Nashville, TN 37232, USA; 102. Vanderbilt Genetics Institute Institute, Vanderbilt University School of Medicine, Nashville, TN 37232, USA; 103. Department of Psychiatry and Behavioural Neurosciences, Offord Centre for Child Studies, McMaster University, Hamilton, ON L8P 0A1, Canada; 104. National Institute of Mental Health, National Institutes of Health, Bethesda, MD 20892, USA; 105. Howard Hughes Medical Institute and Division of Genetics and Genomics, Boston Children’s Hospital, Boston, MA 02115, USA; 106. and Departments of Neurology and Pediatrics, Harvard Medical School, Boston, MA 02115, USA; 107. Laboratory of Genetics, University of Wisconsin-Madison, Madison, WI 53706, USA; 108. Institute for Neurodegenerative Diseases, UCSF Weill Institute for Neurosciences, University of California San Francisco, San Francisco, CA 94143, USA; 109. Quantitative Biosciences Institute (QBI), University of California, San Francisco, San Francisco, CA 94143, USA; 110. Division of Genetics, Boston Children's Hospital, Boston, MA 02115, USA; 111. National Centre for Register-Based Research, Aarhus University, 8210 Aarhus, Denmark; 112. Centre for Integrated Register-based Research, Aarhus University, 8210 Aarhus, Denmark; 113. Department of Biomedicine, Aarhus University, 8000 Aarhus, Denmark.; 114. Centre for integrative Sequencing (iSEQ), Aarhus University, Aarhus, Denmark; 115. Danish Centre for Neonatal Screening, Department for Congenital Disorders, Statens Serum Institut, Copenhagen, Denmark; 116. Institute of Biological Psychiatry, MHC Sct. Hans, Mental Health Services Copenhagen, 4000 Roskilde, Denmark; 117. Genomics plc, Oxford, UK; 118. MRC Centre for Neuropsychiatric Genetics & Genomics, School of Medicine, Cardiff University, Cardiff, UK; 119. Department of Psychiatry, Psychosomatics and Psychotherapy, Center of Mental Health, University Hospital W√ºrzburg, W√ºrzburg, Germany; 120. Department of Clinical Neuroscience, Centre for Psychiatric Research, Karolinska Institutet, Stockholm, Sweden; 121. Psychosis Research Unit, Aarhus University Hospital, 8240 Risskov, Denmark; 122. Mental Health Services in the Capital Region of Denmark, Mental Health Center Copenhagen, University of Copenhagen, 2200 Copenhagen, Denmark; 123. Department of Psychiatry and Psychotherapy, Universit√§tsmedizin Berlin Campus Charit√© Mitte, Berlin, Germany; 124. Department of Family Medicine and Public Health, University of California, San Diego, La Jolla, CA, 92093, USA; 125. Department of Clinical Medicine, University of Copenhagen, 2200 Copenhagen, Denmark

### Supplementary Methods

#### SNV/indel processing

Samples were aggregated from four independent sources: 1) previously published data from the Autism Sequencing Consortium (ASC; total N = 26,268[10,73]); 2) previously published data from the Simons Foundation for Autism Research Initiative (SFARI) Simons Simplex Collection (SSC; total N = 9,170[6,73]); 3) unpublished data from the ASC (N = 5,036); 4) the recently released Simons Powering Autism Research for Knowledge (SPARK initiative; N = 22,766[74]). The distribution of these samples is provided in **Supplementary Table 1**.

| **SNV/ Indel** | ASC Published | ASC New | SSC | SPARK | Total |
|---|---|---|---|---|---|
| Proband | 4,027 | 1,579 | 2,422 | 7,008 | 15,036 |
| Sibling | 347 | 263 | 1,850 | 3,032 | 5,492 |
| Mother | 3,853 | 1,597 | 2,449 | 6,363 | 14,261* |
| Father | 3,853 | 1,597 | 2,449 | 6,363 | 14,261* |
| Case | 5,591 | | | | 5,591 |
| Control | 8,597 | | | | 8,597 |
| Total | 26,268 | 5,036* | 9,170 | 22,766* | **63,237**** |

***Supplementary Table 1****. We aggregated samples from four independent cohorts, including previously published ASC samples, SSC samples, unpublished ASC samples, and SPARK samples. Count summation discrepancies: * Family Utah_1574 is also in SPARK with a different offspring. **1 mother was also a proband in the new ASC data.*

Across these consortia datasets, samples were processed and jointly genotyped in four batches. The first two batches included the published and unpublished ASC and SSC cohorts: 1) ASC B14 - ASC samples through consortium sequencing batch 14 plus the SSC (N = 24,099, 4,632 new), 2) ASC B15-16 - ASC batches 15 and 16 (N = 832, all new). The latter two batches included two independent releases of the **SPARK** cohort: 3) SPARK Pilot initial release (N = 1,379), 4) the SPARK.27k.201909 release (N = 21,387). Finally, raw data was not available for 1,404 and their family members that were reported in Satterstrom *et al.*, and these variants were lifted over directly to GrCH38 (**Supplementary Table 1**).

| SNV/ Indel | ASC B14 SSC | ASC B15_B16 | SPARK Pilot | SPARK | Satterstrom *et al*. Lifted Over | Total |
|---|---|---|---|---|---|---|
| Proband | 7,287 | 283 | 465 | 6,543 | 458 | 15,036 |
| Sibling | 2,348 | 11 | 0 | 3,032 | 101 | 5,492 |
| Mother | 7,233 | 269 | 457 | 5,906 | 397 | 14,261* |
| Father | 7,232 | 269 | 457 | 5,906 | 398 | 14,261* |
| Case | | | | | 5,591 | 5,591 |
| Control | | | | | 8,597 | 8,597 |
| Total | 24,192 | 832* | 1,379 | 21,385 | 15,542* | 63,327** |

***Supplementary Table 2****. We curated and applied GATK to VCFs for de-duplicated samples in ASC+SSC and SPARK columns for SNVs/indels calling in a uniform pipeline. ASC+SSC totals include the "ASC B14" data[10] and new samples from "ASC B15_B16" VCFs. SPARK totals include the "SPARK Pilot" and "SPARK.27k.201909" VCFs. Count summation discrepancies: * Family Utah_1574 is also in SPARK with a different offspring. ** 1 mother was also a proband in the new ASC data.*

### ASC+SSC

The de-duplicated set of 24,932 samples of the ASC+SSC cohort was processed identically, with raw sequencing outputs aligned to the GRCh38 reference genome and variants jointly called using GATK[75]. Briefly, samples were called individually using local realignment by GATK HaplotypeCaller in gVCF mode, such that every position in the genome is assigned likelihoods for discovered variants or for the reference. Individual gVCF outputs were then post-processed, assigning "blocks" of homozygous reference calls one of three qualities: no coverage/evidence, genotype quality < Q20, or genotype quality >= Q20. These compressed per-sample gVCF genotype data, alleles, and sequence-based annotations were then merged using GenomicsDB (https://github.com/Intel-HLS/GenomicsDB), a datastore designed for genomics that takes advantage of a cohort's sparsity of variant genotypes. Samples were jointly genotyped for high confidence alleles using GenotypeGVCFs on all autosomes. Variant call accuracy was estimated using Variant Quality Score Recalibration (VQSR) as in the GATK Best Practices.

### SPARK

For the SPARK Pilot, we accessed GRCh38-aligned bam files for 1,379 samples from SFARI via the /SPARK/Pilot/BAM/ directory in the Globus file sharing platform. The bam files were subsequently realigned to the GRCh38 reference and fed into the same GATK pipeline as outlined above for the ASC+SSC data. For the SPARK main freeze, 27,270 individual GATK-produced gVCFs were downloaded from SFARI via the /SPARK/Regeneron/SPARK_Freeze_20190912/Variants/GATK/ directory in Globus. Per provided documentation, the gVCFs were generated with GATK v4.1.2.0 HaplotypeCaller with default thresholds for calling, using the GRCh38 reference and target files provided by Regeneron (genome.hg38rg.fa & xgen_plus_spikein.b38.bed, respectively). New quality scores, lenient processing of VCF files, and 100bp padding for intervals were also used. We carried out subsequent joint calling of all 27,270 sample gVCFs via GATK to produce one unified VCF.

#### Previously published variants

*De novo* variants from 559 samples were directly carried over from Satterstrom *et al.*; they had been curated from a number of previously published studies[5–9]. The genes that each variant impacted as well as the type of impact (PTV, missense, etc) were retained.

### SNV/indel filtering

#### Creation of working datasets

VCF processing was carried out in Hail 0.2 (https://hail.is) to create a working dataset for each of the four batches of samples. After VCF import, multi-allelic sites were split and variants were annotated using the Variant Effect Predictor (VEP)[76]. Hail's ibd() function was used to verify reported pedigrees and check for duplicate samples within and across datasets. Annotations such as gene and functional consequence were assigned to each variant by prioritizing coding canonical transcripts in the VEP output. Low-complexity regions (using https://github.com/lh3/varcmp/blob/master/scripts/LCR-hs38.bed.gz) were removed. Sexes were imputed with the impute_sex() function in Hail, after which several genotype filters were applied.

Genotypes were filtered to remove calls with a depth below 10 (except for male hemizygous regions, where calls were removed if depth was below 7) or above 1000. Homozygous reference calls were filtered if the genotype quality (GQ) was below 25, while heterozygous and homozygous variant calls were filtered if the phred-scaled likelihood of the call being homozygous reference (PL[HomRef]) was below 25. Additionally, heterozygous calls were dropped if they were in a hemizygous region in a male sample, and any call apparently on the Y chromosome in a female sample was dropped.

Genotypes were further filtered if the allele balance (that is, the number of reads supporting the alternate allele divided by the depth) of a heterozygous call was below 0.25; if the probability of the allele balance (based on a binomial distribution with mean 0.5) was below 1e-9; or if the number of informative reads supporting a heterozygous call (counting reads supporting either the reference or alternate allele) or homozygous call (counting reads supporting the alternate allele) was less than 90% of the depth. Variants were then dropped if they had a call rate below 10% or a Hardy-Weinberg p value less than $10^{-12}$, and a working dataset was written.

### *De novo* variant calling and quality control

To call *de novo* variants from the working datasets, a filter requiring a GQ of at least 25 was applied to every genotype, and Hail's de_novo() function was called with variant frequencies from the non-neuro subset of gnomAD GRCh38 exomes v2.1.1 (gs://gnomad-public/release/2.1.1/liftover_grch38/ht/exomes/gnomad.exomes.r2.1.1.sites.liftover_grch38.ht) used as priors. After calling, putative *de novo* variants were dropped if they were present within this gnomAD subset at a frequency greater than 0.1%, or if they were present within their own dataset at a frequency greater than 0.1%. Variants were also dropped if they contained "ExcessHet" in the Filters field, had a proband allele balance of less than 0.3, or had a depth ratio (child read depth divided by the sum of parental read depth) of less than 0.3.

The following filtering steps were dataset-specific. For the ASC v17 dataset, variants were kept only if the calling algorithm marked them as "HIGH" or "MEDIUM" confidence, with the medium-confidence calls limited to a maximum allele count in the dataset of 3. SNPs with a VQSLOD below -20 were dropped, as were indels with a VQSLOD below -2. The proband allele balance threshold was raised to 0.4 for variants from cell line-derived samples, and variants that appeared more than three times in the remaining set were dropped (this dropped two different synonymous variants in OR13C2). For the ASC B15-B16 dataset, only "HIGH" confidence variants with a VQSLOD at least -3.2 were kept. The call rate threshold was raised to 0.9, and one call was dropped that had reads supporting the alternate allele in a parent's homozygous reference genotype. No variant appeared more than once in the remaining set. For the SPARK Pilot dataset, only "HIGH" confidence variants were kept. SNPs were dropped if they fell in a VQSR tranche above 99.5, and indels were dropped if they fell in a VQSR tranche above 96.0. As with the B15-B16 dataset, the call rate threshold was raised to 0.9, one call was dropped that had reads supporting the alternate allele in a parent's homozygous reference genotype, and no variant appeared more than once in the remaining set. For the SPARK main freeze, only "HIGH" or "MEDIUM" confidence variants were kept, with the medium-confidence calls limited to a maximum allele count in the dataset of 3. Variants with a VQSLOD below -11.6 were dropped. The call rate threshold was raised to 0.5, and variants that appeared more than three times in the remaining set were dropped (this dropped one intronic variant in THBS2 and another in HLA-A).

Following these dataset-specific steps, one variant was selected per person per gene, prioritizing variants with more severe consequences. Finally, for each dataset, samples were dropped if their count of coding *de novo* variants was significantly greater (i.e. $p < 0.05/n$) than expected based on a Poisson distribution with the dataset's observed mean number of coding variants per sample (in ASC v17, this dropped 28 samples with more than 9 coding *de novo* variants; in both ASC B15-16 and the SPARK Pilot, this would have dropped any samples with more than 7, but there were none; in the SPARK main freeze, this dropped 3 samples with more than 9).

### Case-control variants

ASC case-control samples consisted of Danish iPSYCH samples and Swedish PAGES samples. Rare variant counts for 4,863 autism and 5,002 control samples from the iPSYCH cohort were available from the supplement of Satterstrom *et al.*, where rare variants were defined as those with an allele count no greater than 5 in the combination of the iPSYCH data with non-Finnish Europeans from the non-psychiatric subset of gnomAD (a total of 58,121 people). In addition to samples labeled as "Autism", samples labeled as "Both" in that study (meaning that an individual had both autism and ADHD diagnoses) were used as autism cases for our purposes.

Rare variant counts for 728 autism and 3,595 control samples from the PAGES cohort were taken from Satterstrom *et al.*, where rare variants were defined as those with an allele count no greater than 5 in the 18,153 combined parents, cases, and controls in the dataset, as well as an allele count no greater than 5 in the non-psychiatric subset of ExAC r0.3 (45,376 people). Counts were removed for 17 cases for whom parental sequences became available, so that they are now included in our family-based data instead.

### Inherited variants

Counts of transmitted and non-transmitted alleles were produced starting from each of the four working datasets described above. First, variants were dropped that had been marked "ExcessHet" in the Filters field by GATK or had allele frequencies greater than 0.1% in either their own dataset or the non-neuro subset of gnomAD GRCh38 exomes v2.1.1. In addition, a filter requiring a GQ of at least 25 was applied to every genotype. Hail's transmission_disequilibrium_test() function was then called to count transmitted and untransmitted alleles for each variant in family-based data.

From these raw results, dataset-specific filters were applied: VQSLOD thresholds required variants to possess a VQSLOD value of at least 5.13 in the ASC v17 dataset, 24.12 in the ASC B15-B16 dataset, and 6.01 in the SPARK main freeze. In the SPARK Pilot, all variants that did not pass VQSR were dropped. These thresholds were based on identifying the VQSLOD values necessary to balance the transmission/non-transmission of singleton synonymous SNVs from the parents or the transmission/non-transmission of non-coding indels, and selecting the more stringent VQSLOD value. For the ASC v17 dataset in particular, additional filters were applied to standardize data from samples originally sequenced across a span of several years. Specifically, variants were retained only if they possessed a strand odds ratio no greater than 3, a Read Position Rank Sum of at least -8, and a quality by depth of at least 1 (for SNVs) or 3 (for indels). After application of these filters, final counts of transmitted and non-transmitted alleles were produced for variants of different classes (e.g., PTV, MisB, MisA).

### CNV processing

For the subset of samples with available raw genomic data (**Supplementary Table 3**), we employed GATK-gCNV for exome CNV detection, along with an additional supplement of 7,832 general research use (GRU) controls. GATK-gCNV is specifically designed to adjust for known bias factors of exome capture and sequencing (e.g GC content), while automatically controlling for other technical and systematic differences. Briefly, raw sequencing files were compressed into read counts over the set of annotated exons and used as input, and a PCA-based approach (see below) was implemented on observed read counts to distinguish differences in capture kits (**Supplementary Fig. 1**), followed by a hybrid density and distance based clustering approach to curate batches of samples for parallel processing. Filtering metrics derived from the underlying Bayesian model were included for each detected variant, and were then tuned to balance between sensitivity and specificity.

| **CNV** | ASC<br>SSC | SPARK | GRU<br>Control | Total |
|---|---|---|---|---|
| Proband | 6,827 | 6,867 | | 13,964 |
| Sibling | 2,024 | 2,983 | | 5,007 |
| Parents | 12,600 | 13,803 | | 26,403 |
| Case | 608 | | | 608 |
| Control | 3,480 | | 7,832 | 11,312 |
| Total | 25,539 | 23,653 | 7,832 | 57,294 |

***Supplementary Table 3****. Number of samples for each cohort that passed our exome CNV calling pipeline quality control.*

#### Cluster curation by PCA analysis

A set of 7,981 target regions were chosen from among 7 popular exome enrichment kits (Agilent_V4, Agilent_V5, Agilent_V6r2, Agilent_V7, NimbleGen SeqCap EZ2, NimbleGen SeqCap EZ3, Illumina TruSeq), such that each chosen region is uniquely targeted by 1 of the 7 kits. Depth of coverage was collected over the 7,981 regions for all samples, and a primary PCA analysis was conducted to identify samples that were prepared with different enrichment kits (**Supplementary Fig. 1**). After kit determination, a secondary PCA analysis was conducted for samples on the same enrichment kits to determine final cluster assignment via a hierarchical clustering approach.

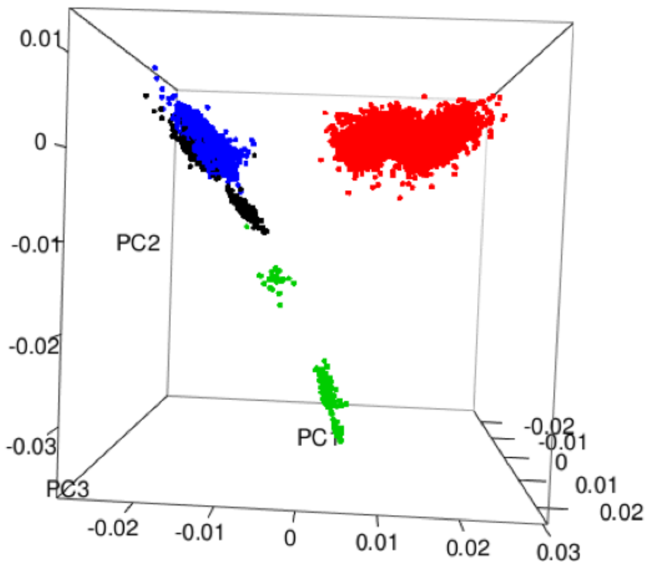


***Supplementary Fig. 1***. *PCA of read counts from exome sequencing data of 7,981 specially chosen regions. Clear capture kit and technical artifacts can be observed. The first 3 PCs of the chosen target regions clearly differentiate the capture kits used to enrich each sample. Red corresponds to Illumina, black Agilent, blue Agilent 1.1, and green with Nimblegen. Secondary clustering in 3d space of each of the determined capture clusters was carried out to create batches for GATK-GCNV to process for CNVs.*

### Curation of bins and exons modeled

Due to the differential exon targeting and differences in transcriptome annotation, we standardized the set of intervals/bins/exons over which GATK-gCNV queried for CNV events to better homogenize our cohort. To construct this list of bins, we took all exons from the Gencode V33 annotation and collapsed them to non-overlapping intervals. Collapsed intervals longer than 800bp were evenly split into subdivisions less than 1,600bp, in an attempt to increase the number of intervals/data points for GATK-gCNV to identify copy number events. The resulting intervals were padded by 100 basepairs in a non-overlapping, equal manner. Intervals over which <50% of samples were covered by 10 reads, >50% segmental duplication content, < 90% mappability, or GC content outside of the interval (0.1, 0.9), were excluded from further processing.

### Cluster processing with GATK-gCNV

GATK-gCNV can be run in 2 modes: cohort, case. Cohort mode takes the set of input samples, constructs a model on the copy number states of each input region that passes filtering metrics, and subsequently makes determinations of the copy number events in those samples. Case mode takes a pre-computed model output from cohort mode, and proceeds to directly determine copy number events in a set of supplied samples, offering time and cost savings if the samples are similar enough. Cohort mode was run for every cluster determined by PCA analysis, using 200 samples per cluster. As most PCA-defined clusters consisted of more than the 200 samples used to create the model in GATK-gCNV cohort mode, the remaining samples from each cohort were analyzed with GATK-gCNV using case mode with their respective cohort mode models.

GATK-gCNV parameters included:
num_interval_scatter = 12500
gcnv_interval_psi_scale = 0.01
gcnv_max_bias_factors = 6
gcnv_p_active = 0.1
gcnv_o_alt = 0.0005
gcnv_sample_psi_scale = 0.01

### Variant compilation, quality control, and annotation

For each sample processed using GATK-gCNV, we extracted non-reference copy number events from the output. All calls with quality score (QS) >= 20 were considered for defragmentation (concatenation). Each candidate call was translated onto the interval-space and extended by 50% in width on both ends; calls that had consistent copy number and overlap when extended were merged into a single call.

All calls for samples and clusters were compiled into a single table. Then single-linkage clustering requiring 80% reciprocal overlap on the interval-space was used to determine when multiple calls were the same variant. Site frequencies were assigned to each variant as the proportion of samples that had either a deletion or duplication of this variant in both an overall and cluster-specific manner. We annotated the number of callable exons that each variant overlaps by counting the number of exons from Gencode V33 protein-coding canonical transcripts that passed filtering during the bin curation process detailed above.

For quality control of variants, we used sample, variant, and call-level metrics. Variants were retained if the site-frequency of the variant was rare (less than 1%), spanned 3 or more callable exons, and less than 50% of the intervals that compose the variant had maximum cluster-specific site frequency greater than 2.5%. Individual call-level filters required that homozygous or copy number 0 deletions attain QS >= 400, heterozygous or copy number 1 deletions had QS >= 100, or duplications had QS >= 50. For sample-level quality control, samples passed if their number of raw CNV calls detected by GATK-gCNV did not exceed 200 and the number of calls with QS>=20 did not exceed 35.

We annotated a deletion to impact a gene if >=10% of the non-redundant exon-basepairs were overlapped by the deletion; for a duplication, it impacted a gene if >=75% of the non-redundant exon-basepairs were overlapped by the duplication. CNVs were annotated against a list of 75 curated genomic disorder (GD) loci (**Supplementary Table 4.8**). A call was considered a genomic disorder CNV if it shared 50% reciprocal overlap with the annotated GD.

Familial CNV delineation

For samples from a family fully characterized for CNVs (all 3 members), we further determined inheritance and *de novo* status of CNVs. High-quality CNVs in the offspring for which more than 30% of the callable exons overlapped a called CNV from either parent were deemed inherited. CNVs that had less than 30% of their intervals in such matching calls underwent secondary screening. They were considered *de novo* only if three criteria were met: average normalized copy number was within 0.3 of the reported copy number; if the minimum normalized copy difference between the child and either parent was > 0.7; and less than 50% of the normalized copy number of the constituent intervals had median absolute deviation across samples greater than 0.5. Variants were considered inherited from a mosaic parent if the normalized copy number of the parents across that site shows skew towards the offspring's copy number. The threshold for the skew was determined as a function of the number of intervals the CNV spans.

Gamete-of-origin determination

*De novo* CNVs were evaluated for gamete-of-origin where possible using the SNP/indel joint-called VCF. For each *de novo* deletion in the child, the region was queried for variants in the offspring and both parents. For each variant in the child within that region that is either 0/1 or 1/1, we recorded which parent had the corresponding variant and which had the reference. For *de novo* duplications, we recorded variants for which the child was 0/1 and had B-allele frequency > 60%. For those variants, we then recorded if one parent was homozygous reference, while the other parent harbored the 0/1. A binomial test of the number of variants that are consistent with the mother versus the father was carried out for binomial parameter p=0.5. Rejection of equality by this test allows assignment of the variant's origin to the parent with the higher number of consistent variants.

### CNV Benchmarking

We had access to 7,165 samples for which matching genome (WGS) and exome sequencing data were available for benchmarking comparisons. The ground truth data was considered to be the CNVs called from WGS using the ensemble machine learning method GATK-SV[47].
For GATK-gCNV, we benchmarked the performance of the full 7,165 samples against the WGS ground truth (**Supplementary Figs. 2, 3**). Sensitivity was measured by the proportion of CNVs called from WGS data that have a match in the GATK-gCNV callset. Specifically, for each variant, if at least 75% of the samples that have that CNV in the WGS data also had a GATK-gCNV CNV call with a consistent direction (deletion or duplication) that overlapped at least 30% of the callable exons, this was considered a success. For CNVs called by GATK-gCNV, their positive predictive value (PPV) was measured by requiring that 75% of the GATK-gCNV samples with that call have a match to the WGS calls (ground truth) with at least 30% exon overlap.

We also benchmarked two other popular exome CNV calling methods, XHMM[77] and CoNIFER[78] in comparison to GATK gCNV (**Supplementary Fig. 2**) and found that GATK gCNV achieved the highest sensitivity and PPV relative to WGS ground truth.

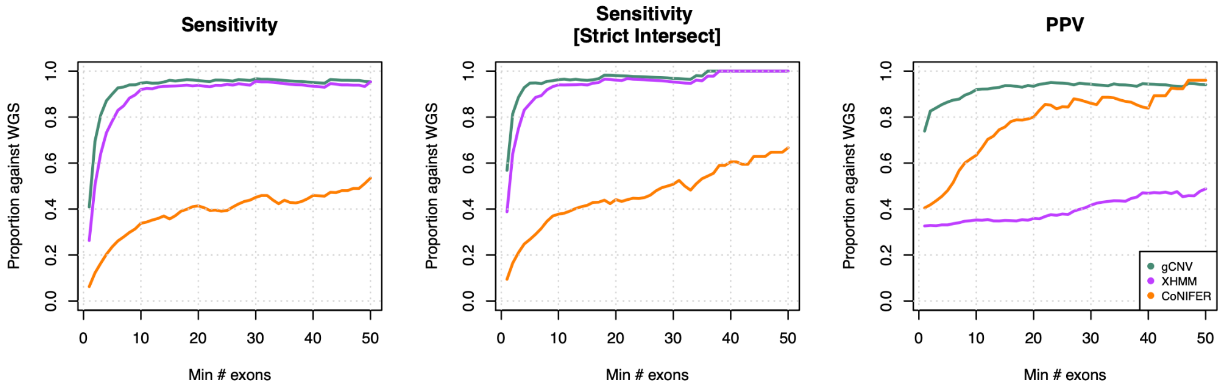


***Supplementary Fig. 2.*** *For comparison of exome CNV methods, we benchmarked approximately 1,000 samples with GATK-gCNV, XHMM, and CoNIFER, following available best practices for each of the respective methods. Sensitivity and PPV evaluation criteria directly mirror those described above for comparing GATK-gCNV to the ground truth, with substitution of XHMM and CoNIFER for GATK-gCNV as appropriate. Sensitivity of each exome CNV method to capture the ground truth genome calls were measured as a function of the exon sizes through 2 different exon sets: the callable exon set dictated by each caller, and the intersection of all 3 call-able exon sets. PPV for each method is measured as a function of callable exons reported by each method. GATK-gCNV greatly outperformed the other methods, achieving higher sensitivity while maintaining greater PPV.*

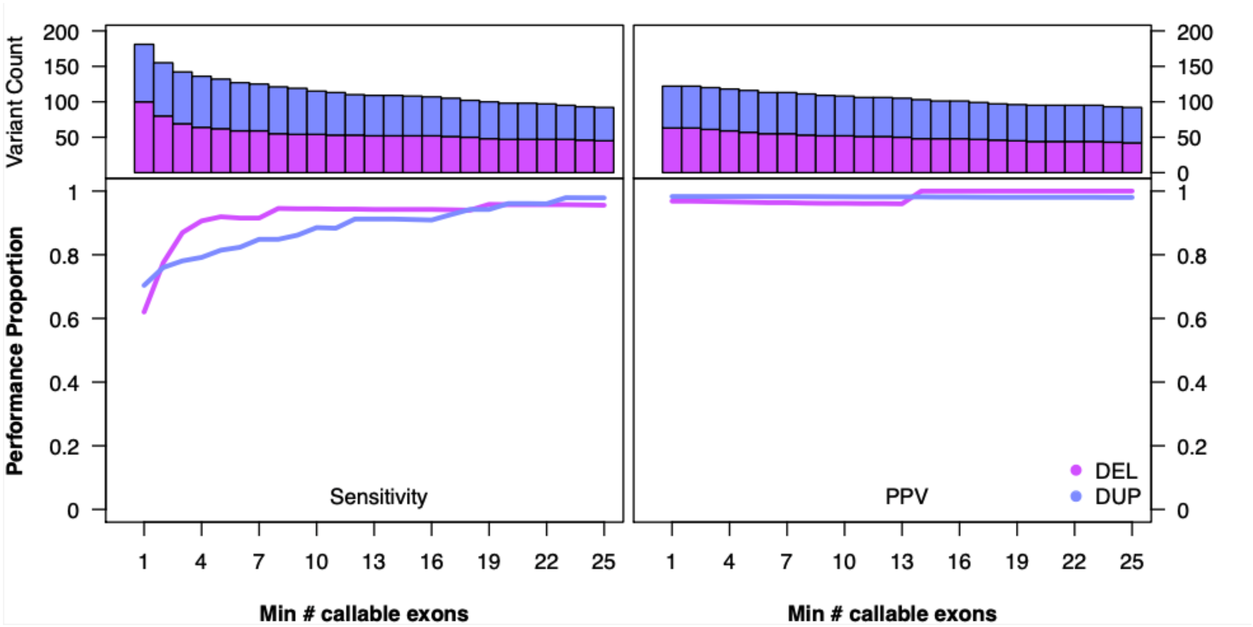


***Supplementary Fig. 3.*** *Using a set of 7,165 samples for which we have matching WES data and gold-standard WGS CNV calls[47], we benchmarked the performance of CNVs derived from WES data using GATK-gCNV as a function of the number of callable exon of canonical transcripts. (A) Sensitivity: using rare WGS CNVs (site frequency < 1%) as ground truth, we find that GATK-gCNV achieves 87% and 78% sensitivity at recapitulating gold-standard WGS calls for rare deletions and duplications that span 3 or more such exons. (B) PPV: given rare WES CNVs (site frequency < 1%), we looked for corroborating calls from the gold-standard WGS callset. We find that GATK-gCNV achieves 97% and 98% PPV for WES deletions and duplications that span 3 or more exons.*

### TADA Bayesian Framework for Gene Association

TADA is a Bayesian framework that produces gene-level measures of evidence for association that can be transformed into a false discovery rate[41]. Broadly speaking, for a given variant type at gene, TADA produces a Bayes Factor (BF) to measure statistical evidence, taking as input the count of variant events, mutation rate, number of samples, and a prior on the risk of a variant in this gene. BF can be readily combined across different variant types for the same gene through multiplication, to arrive at a total measure of association for a given gene. This total BF can then be directly transformed into a FDR and the appropriate statistical threshold can be applied to extract a candidate gene list. In the previous TADA study[10], evidence was aggregated for *de novo* PTV, misB, and misA variants, as well as case/control PTVs to find 102 genes significant at the FDR < 0.1 threshold.

### Modes of evidence included

We have expanded evidence integration to all of the following combinations, comprising a full 5x3 combination of variant types by inheritance classes (PTV, MisB, MisA, DEL, DUP) x (*de novo*, case-control, inherited).

### Preparing CNVs for TADA Integration

Due to the challenge in pinpointing driver genes within large, recurrent CNVs, we opted to exclude NAHR-mediated CNVs and focus on modeling smaller CNVs that impact 8 or fewer constrained genes (defined by LOEUF < 0.6).

### Prior Elicitation for TADA using Empirical Bayes

**PTVs**

We used an empirical Bayes approach, borrowing information across genes, to estimate the prior relative risk (gamma) for ASD association of each gene. This consists of combining 2 components: (1) calculate the relative risk of PTVs for each inheritance type and (2) the estimated fraction of genes that substantially influence risk ASD. Gamma for each gene is then calculated as (1) divided by (2), smoothed over a measure of constraint. In Satterstrom *et al. Cell* 2020, the smoothing is conducted over pLI, which is now superseded by the updated loss-of-function observed/expected upper bound fraction (LOEUF) [39]. In [10], the fraction of genes that substantially influence risk ASD is assumed to be constant over the entire pLI range. We relaxed this assumption because ASD associated genes are heavily concentrated among the constrained rather than the unconstrained genes. We used equations in the supplement of [41] to estimate the proportion of genes that are risk genes, specifically methods of moments estimation for the proportion of risk genes by solving for the following equations:

$$(\bar{\gamma} - 1)k = \frac{C}{2N\bar{\mu}} - m. \tag{29}$$

$$k\mathrm{E}\left(p|\bar{\gamma}, \beta\right) + (m - k)\mathrm{E}\left(p|\gamma = 1\right) = M, \tag{33}$$

*C* = the observed number of LOF events in all genes
*M* = the number of genes with more than 1 LOF event
*m* = the number of genes
*N* = the number of probands
*k* = the number of ASD genes
$\gamma$ = the relative risk

(33) is [number of multi-hit genes under alt] + [number of multi-hit genes under risk=1]

Using equations (29) and (33) and by using a sliding window of 20% of the genes, ordered by LOEUF, we computed a rolling-average estimate for the fraction of risk genes (**Supplementary Fig. 4**).

For the *de novo* prior, (1) is estimated as the relative enrichment of variants in affected versus unaffected offspring; for case-control prior, (1) is estimated as the relative enrichment of variants in cases versus controls; while for the inherited prior, (1) is estimated as the relative enrichment of transmitted versus untransmitted variants. The estimated prior relative risks all decrease with lower genic constraint as expected, with *de novo* prior being the strongest, followed by case-control then inherited respectively. We averaged the priors between our ASC/SSC cohort and the SPARK cohort.

**Missense variants**

Similar to the approach in Satterstrom *et al.*[10], the prior relative risks of *de novo* misA and misB variants, separately, were estimated as (R-1)/0.05+1, where R is the enrichment ratio of the variant class, and 0.05 is the proportion of risk genes across the genome. We adopted the same approach. For *de novo*, R is the ratio of observed counts from affected versus unaffected offspring; for case-control, R is the ratio of observed counts from cases versus controls; and for inherited, R is the ratio of observed counts of transmitted to untransmitted variants from parents to their affected offspring. All priors are floored at 1.05x relative risk.

**CNVs**

To estimate the prior relative risk of *de novo* deletion CNVs, we use the estimated prior risk for *de novo* PTVs falling in constrained genes within the CNV. Specifically, the estimated prior risk of a CNV is the summation of the estimated PTV prior risk for constrained genes that are annotated for that CNV. For *de novo* duplications, we first computed the same sum, then downweighted the sum proportional to the observed ratio of *de novo* deletions/duplications. This accounted for decreased penetrance of duplications compared to deletions. For inherited and case/control CNVs, we followed the same approach, substituting the corresponding prior PTV risk estimate.

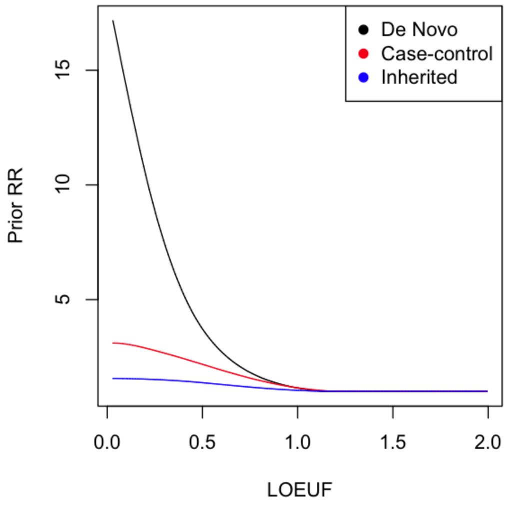


***Supplementary Fig. 4.*** *Based on our prior elicitation procedure, we computed separate priors for the risk of de novo, case/control, and inherited variants in our ASD dataset. Increasing risk trends strongly with increasing constraint, as measured by LOEUF; while de novo variants represent more risk compared to case/control, and inherited counterparts given the same constraint.*

### BF calculation using TADA

#### ***De Novo*** **- PTV/MisA/MisB**

For each gene, PTV, MisA, and MisB BFs were calculated per previously described implementation of TADA[10,41], taking into account sample size and using updated mutation rates and prior relative risk. Mutation rates for PTVs were updated using gnomAD (v.2.1.1) released LOF mutation rates, scaled so that the expected number of *de novo* PTVs matched the observed number in unaffected siblings. MisA and MisB mutation rates were carried over from Satterstrom *et al.*, and genes previously missing MisA or MisB mutation rates were given estimates using gnomAD (v.2.1.1) missense mutation rate scaled by the average missense to MisA or MisB mutation ratio respectively. MisA and MisB mutation rates were scaled so that expected and observed *de novo* counts matched that of unaffected siblings.

BFs for *de novo* PTV/MisA/MisB variant types were calculated separately for our ASC/SSC sub-cohort and the SPARK cohort, to facilitate direct comparisons to *de novo* PTV/MisA/MisB evidence from the DDD study.

#### ***De Novo*** **- CNVs**

BFs were estimated per CNV, analogously to the per gene calculation of BFs for PTVs/MisA/MisB variants and separately for deletions and duplications. We excluded from modeling CNVs that were NAHR-mediated GD events, and focused on the contribution of constrained genes within CNV segments. Mutation rates of deletions and duplications were estimated using data from the unaffected siblings, where the rate was approximated as the number of observed *de novo* deletions or duplications that span X constrained genes, divided by the total number of sequences of X constrained genes. Given the prior relative risk and the mutation rate, each unique CNV had a total BF estimate, which was then distributed to the constituent constrained genes relative to their LOEUF measure. For example, if a CNV had a total BF=10 and it spanned two constrained genes, A and B with LOEUF 0.1 and 0.4 respectively, we distributed BF=8 to gene A and BF=2 to gene B, consistent with the 4:1 relative constraint ratio estimated by LOEUF.

**Case/control - PTV/MisA/MisB**
For each gene, PTV, MisA, and MisB BFs were calculated per previously described implementation of TADA[10,41], taking into account case/control sample sizes and updated priors for case/control variants.

**Case/control - CNV**
Like *de novo* CNVs, BFs were estimated per CNV using the case/control PTV/MisA/MisB framework described previously, with prior relative risk estimated as the sum of the prior case/control PTV relative risk of the constituent constrained genes. For duplications in this setting, we also used a downweighting factor for prior relative risk compared to a deletion of the same gene, analogous to the *de novo* CNV setting. The BF for each CNV was then distributed to the genes that constitute the CNV, proportional to LOEUF, as in the *de novo* setting.

**Inherited - PTV/MisA/MisB**
For each gene, PTV, MisA, and MisB BFs were calculated per previously described implementation of TADA for case/control data[10,41], using updated priors for inherited variants as described above, and using a sample size of all affected offspring in trio settings.

**Inherited - CNV**
For inherited CNVs, BFs are estimated per CNV, using the case/control PTV/MisA/MisB framework described previously, with prior relative risk estimated as the sum of the prior inherited PTV relative risk of the constituent constrained genes. Duplications also used a downweighting factor for prior relative risk compared to a deletion of the same gene, analogous to the *de novo* CNV setting. The BF for each CNV was then distributed to the genes that constitute the CNV, proportional to LOEUF.

### BF integration using TADA

For each variant class (PTV, MisB, MisA, deletion, and duplications), we set a floor to the BF of 1, so that evidence against association based on one variant type does not degrade evidence for association of another variant type. For example, in some genes, PTVs dominate the signal for association and they are somewhat or strongly depauperate in signal from missense variation; for others, the pattern is reversed. We believe such patterns trace to how allelic architecture relates to risk for ASD. Setting a floor for the BF preserves signals related to allelic architecture.

Within each variant class, however, we allowed the BF evidence to offset each other when appropriate. For instance, BF > 1 in unaffected siblings, arising from non-zero *de novo* counts in siblings, are used to offset the BF of that same gene in affected siblings. Operationally, this is achieved by calculating the BF for a certain gene contributed by PTVs as: BF_PTV = floor(BF_PTV_probands / BF_PTV_siblings, 1).

Finally, the total gene-level BF was calculated as the product of the BFs across the 5 variant classes: BF = BF_PTV * BF_MisA * BF_MisB * BF_deletion * BF_duplication. Of note, to ensure no gene was nominated from CNV evidence alone, we set BF_deletion and BF_duplication to 1 if the total BF evidence of PTV/MisA/MisB did not exceed 5.

All BFs are subsequently converted to a posterior probability, which is in turn used to compute a FDR to nominate genes of relevance.

### Applying TADA to DD data

We accessed the summary tables released by the DDD in Kaplanis *et al*., detailing de novo variants detected and gene-level variant counts in 31,058 trios where the offspring is diagnosed with developmental disorders. To calculate the number of PTVs per gene, we aggregated Kaplanis *et al.* released variants with annotated consequences of ("frameshift_variant", "splice_donor_variant", "splice_acceptor_variant", "stop_gained"). For synonymous counts, we aggregated ("synonymous_variant", "stop_retained_variant")-labeled variants. We annotated missense variants ("missense_variant") with MPC scores, and using those MPC scores, we assigned misB and misA status, and aggregated counts per gene.

To create TADA-DD, we supplied the per-gene counts of PTVs, misA, and misB variants to TADA, in the same manner as we supplied our ASD cohort counts. TADA-DD BFs are combined with those from the ASD cohort on a per-gene basis, allowing us to estimate FDR on a combined NDD super-cohort (TADA-NDD).

### Comparison of TADA-DD and denovoWEST from Kaplanis *et al.*

Kaplanis *et al.* reports 19,654 genes, of which 285 are significant at an exome-wide threshold. Of the 18,128 autosomal genes investigated by our study, 17,919 (99%) have a match from Kaplanis *et al,* including all 252 autosomal genes significant in Kaplanis *et al*. 237/252 (94%) of the Kaplanis *et al.* denovoWEST exome-wide significant genes also appear in the TADA-DD FDR ≤ 0.001 list.

We also measured the concordance of the Bayesian TADA-DD FDR with the frequentist denovoWEST estimates of gene significance reported in Kaplanis *et al.* by transforming the

Kaplanis p-values (denovoWEST_p_full) into FDRs (FDR denovoWEST) using the R function p.adjust(method=”fdr”). A pairwise plot of TADA-DD FDR with transformed Kaplanis FDR reveals high concordance (cor=0.95) on the log scale, signaling convergence in evaluation of gene-level evidence between our studies, and allowing us to integrate the Kaplanis variant data in our Bayesian framework.

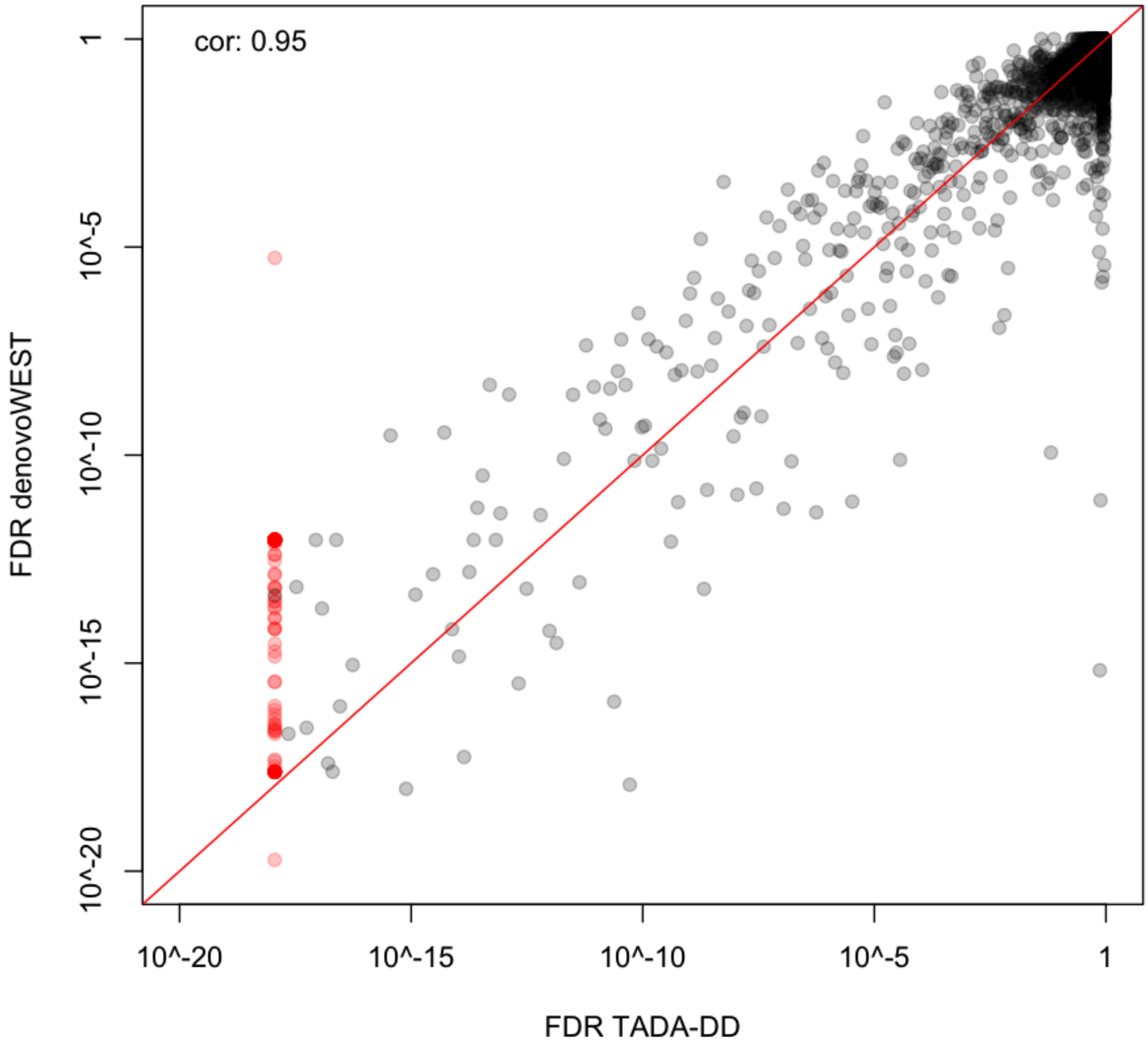


***Supplementary Fig. 5.*** *By using the provided per-gene p-values supplied by Kaplanis et al. (denovoWEST_p_full), we can transform the p-values into a FDR (FDR denovoWEST). Comparing the FDRs from TADA-DD and denovoWEST yields strong concordance between significance measures from the two approaches, attaining a correlation coefficient of 0.95 on the log10 scale. The points in red are genes for which their TADA-DD BF evidence of association is so large that they exceed computational precision, and their FDR computationally returned 0. For these genes, the smallest, non-zero TADA-DD FDR was substituted as a proxy for this figure.*

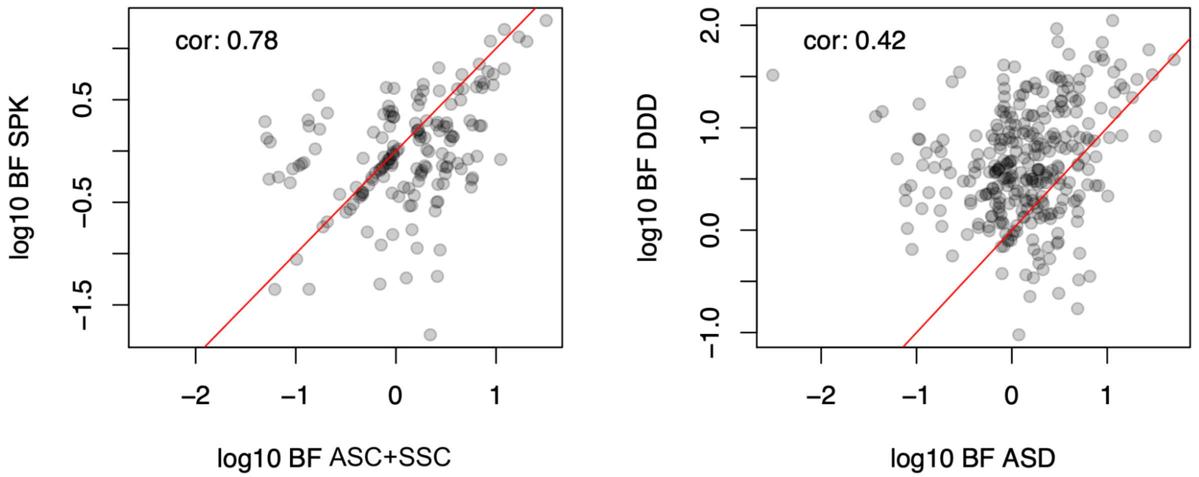


***Supplementary Fig. 6.*** *For the set of 373 TADA-NDD genes with FDR ≤ 0.001, we also examined the concordance of BF evidence between the DD cohort and the subcomponents of the ASD cohort. The ASD cohort is composed of two halves of roughly equal sample sizes - the ASC+SSC portion, and the SPARK portion. We considered only evidence from de novo PTVs, MisB, and MisA variants for calculating BF evidence such that the modes of evidence considered can be identical between the ASD cohort and the DD cohort. The DD cohort was down-sampled to match in size to the total ASD cohort. Pairwise comparison of log10 BF between ASC+SSC and SPARK were plotted for the 373 genes, and achieved a correlation of 0.78; while pairwise comparison of log10 BFs between the combined ASD cohort and the DD cohort only achieved a correlation of 0.42.*

### Cross-validation relation of FDR with relative risk

To relate the estimated FDR to a relative risk measure of *de novo* variants, we conducted cross-validation. We randomly subset the data into 10 equal subsets, taking turns to estimate the TADA model using 9 folds while holding the remaining shard out. For each model fit, we calculated the FDR of each gene, and examined the empirical relative risk of variant types and inheritance class between affected and unaffected individuals for differing FDR thresholds. We repeated this process 1,000 times, randomly assigning samples to the 10 subsets during each iteration. Aggregating over shards and iterations, we estimate that genes with FDR ≤ 0.001 exhibit relative risk of 15-20 for *de novo* PTVs and most damaging missense variants (misB).

### Conditional analysis of cross-cohort association.

For the ASD and DD cohorts, separately, we first converted the set of 18,128 genes q-values into p-values using the following R command: pval = qval * rank(qval) / (max(qval) * length(qval)). Next, we selected genes meeting FDR ≤ 0.05 from the TADA-ASD and TADA-DD cohorts, treating the derived lists separately. For the set of 183 identified TADA-ASD genes, we evaluated the distribution of their p-values from the TADA-DD cohort using the q-value function in R [79] to estimate pi0 and pi1=1-pi0, which is the estimated fraction of the number of genes associated in the DD cohort. (pi0 is the estimated fraction of genes that have no association and for which their p-values would be uniformly distributed on the interval 0-1.) We then did the converse: choosing the set of 477 identified TADA-DD genes, we evaluated the distribution of their p-values from the TADA-ASD cohort to estimate pi1.

ASD-DDD heterogeneity analysis

We analyzed the ASD and DD cohorts separately using TADA with a goal of assessing the nature of heterogeneity between the cohorts. We used only PTV and MisB *de novo* variants to make the data sets strictly comparable. These analyses yielded a total of 464 genes with q-value < 0.05 for at least one cohort; call these the signal genes. Of these, 120 passed the threshold for ASD, 428 for DD and 84 for both cohorts. Tallying PTV and MisB *de novo* mutations per gene, we obtained a two-way contingency table of gene-by-disorder (ASD/DD) for the 464 signal genes. In a previous analysis (Satterstrom *et al*.) of earlier versions of these cohorts, we found highly significant heterogeneity, but this was computed over all genes. Noting that there were now 84 genes significant in both cohorts and therefore each of these genes had a substantial count of mutations in each cohort, we decided to restrict a new heterogeneity analysis solely to those 84 overlapping genes. Even for this set of genes, we observed significant heterogeneity ($X^2$=317.6, DF = 83, p=$3.75x10^{23}$).

Next, we asked which of the signal genes was more tightly connected with either ASD or DD than expected by chance. To do so we formulated an approach that builds on the familiar chi-statistic residual. Before computing the residuals, we needed to overcome the far larger number of mutations present in the DD sample because the standardized residual performs best when the total count of events, per cohort, was equal. To do so, we down-sampled the DD mutations in signal genes to obtain a count of 1001 mutations, which matches the count of mutations in the ASD cohort. This was repeated for 100 repetitions.

**C statistic.** For the C statistic we used a standard log-linear model analysis by conditioning on the row (gene) and column totals (over ASD or DDD). We asked if the residual for ASD was substantially different from that expected under the null. The residual for gene i was defined as

$$C_i^{ds} = \left(dn.asd_i^{obs} - dn.asd_i^{exp}\right)/\sqrt{dn.asd_i^{exp}}$$

where:

$$dn.asd_i^{exp} = dn.asd_i^{obs} \times \left(dn.asd_i^{obs} + dn.ddd_i^{obs}\right)/N$$

with the average over the 100 down-sampling repetitions was recorded as the C-statistic for each gene.

**Mixture modeling**

If genes were independent of cohort, then the C-statistic would be distributed as a standard normal statistic, but this was clearly not true (**Figure 5E**). Genes with unusually few mutations in the ASD cohort produced a negative C statistic and those with unusually many mutations in the ASD cohort produced a positive statistic. Assuming the genes split into two classes, one favoring DD mutations and the other favoring ASD mutations, we fitted a two-component normal mixture model. This calculation was performed using the normalmixEM function in the R library [80]. We restricted the model to have a common standard deviation for both components (option arbvar=F), which was estimated to be 0.527. Although the C statistics varied continuously across the spectrum of values observed, we can estimate the posterior probability a gene is from the DD or ASD component to determine likely group membership. Genes with posterior probability greater than 0.99 for either class were labeled by that class.

**Tree analysis**

To understand in which cell types in early development these genes were expressed, we analyzed two datasets using a new approach called cFIT, the Common Factor Integration and Transfer learning algorithm [66]. cFIT relies on a linear model assuming a common factor matrix shared among datasets, as well as gene-wise location and scale shifts unique to each dataset. It estimates the shared and batch specific parameters through iterative nonnegative matrix factorization and then recovers the batch-free expression for each dataset based on the common factor and factor loadings.

Applying cFIT to fetal cells from two studies, which we will call “Nowakowski”[64] and “Polioudakis”[65], we obtained integrated factor loadings and gene expression for all measured cells (**Supplementary Fig. 7**). The algorithm was applied to a subset of informative genes, including 3,790 genes selected using the Seurat function FindMarkers [81]. To ensure we included all informative ASD and DD risk genes, we also included 676 for which the TADA analysis yielded a q-value < 0.05 for at least one of the three cohorts: ASD, DD, and ASD+DD. After removing genes expressing in less than 5% of cells in either data set, 454 risk genes remained. In total, we identified 3938 genes, which are the only genes included in the downstream analyses.

| Data | Method | # cells post QC | Reads per cell | Total reads | Gene detected per cell | Sample age span |
|---|---|---|---|---|---|---|
| Nowakowski et al 2018 | Fluidigm | 4,261 | 2.2 million | 16 millon | 2403 | 5-37 post conception weeks |
| Polioudakis et al. 2019 | Drop-seq | 33,000 | 52,000 | 118 million | 1049 | 17-18 gestational weeks |

***Supplementary Table 5***. *This table details the data aggregated for our single-cell analyses.*

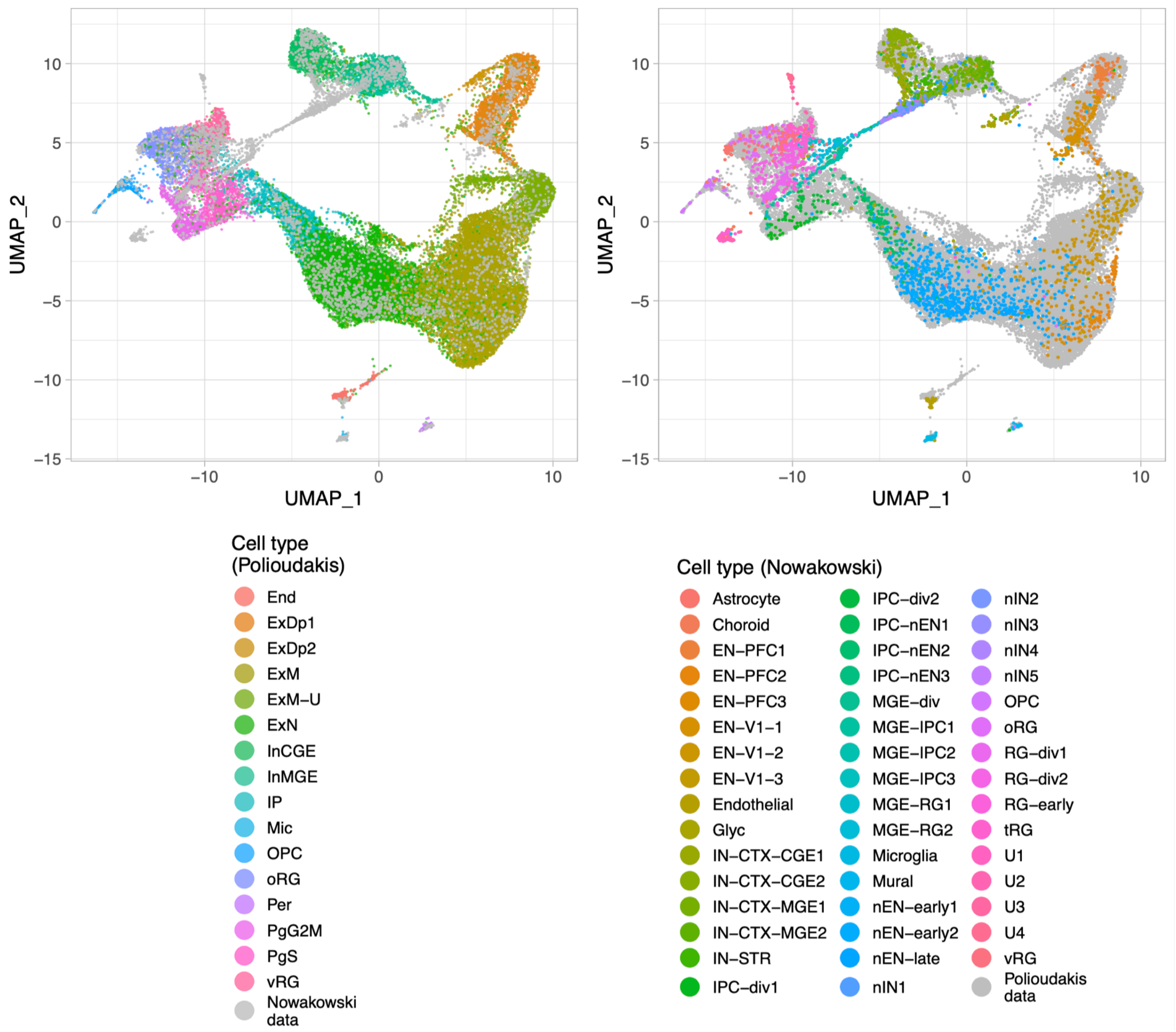


***Supplementary Fig. 7.*** *UMAP of scaled factor loadings obtained from data integration. In Left, only cells from the Polioudakis data are colored into 16 major cell types. Similarly, in Right, only cells from Nowakowski data are colored according to the 48-cell-type label. See SI Appendix, Tabel S2 ([66]) for detailed cell-type annotations from respective studies.*

With the integrated data, we applied unsupervised clustering to identify cell subtypes. While it is typically impossible to definitively determine the right number of clusters, it is clear that cell types should follow a hierarchical structure: major clusters should develop into minor more specialized subtypes. Our MRtree method[67] was developed based on this idea. (**Remark**: MRtree constructs a hierarchical cluster tree from multiresolution clustering to recover levels of specification among cell clusters. The method relies on a stability measure to cut the tree to obtain stable clustering.) To obtain the flat clustering required by MRtree, we used Seurat v4, with resolution varying from 0.05 to 2. By further increasing the resolution, functionally more specialized cell types were separated. After applying the stability criterion to determine the finest stable resolution, the algorithm produced 21 terminal clusters and most exhibited a good mix of cells from each source (**Supplementary Fig. 8a**). The exceptions were two clusters (newborn IN and MGE RG/IPC) derived entirely from Nowakowski cells, which were from a region of the brain not sampled in the Polioudakis study, and one small cluster labeled as ExN, which derived entirely from Polioudakis, which appears to be problematic based on the UMAP (**Supplementary Fig. 8b**). This cluster was removed from downstream analyses. (**Remark**: To avoid the tree growing too large, we excluded undefined types and well separated clusters from tree construction, including 939 cells with missing labels or labeled as U1, U2, U3, U4, Choroid, End, Endothelial, Per, Glyc, Microglia, Mic, and Mural from two studies.)

**Enrichment analysis**

We next performed enrichment analysis for each cluster in the resulting tree to determine if any clusters expressed an unusual number of ASD-predominant or DD-predominant risk genes. Before performing the enrichment analysis, i.e., creating a 2x2 table for expressed gene (yes/no) by risk gene (yes/no), we needed to first identify the set of genes to be included in the analysis, which is defined as the set of genes "expressed" in at least one cell type. Because the integration process often replaces zero values in the gene expression matrix with small positive values, we considered any integrated expression value less than 0.5 to be non-expressed. A gene was considered "expressed" for a particular cell type if its expression was greater than 0.5 for at least 25% of the cells in the terminal clusters. Within this list of genes, we then determined if a gene was expressed or not for a particular cluster.

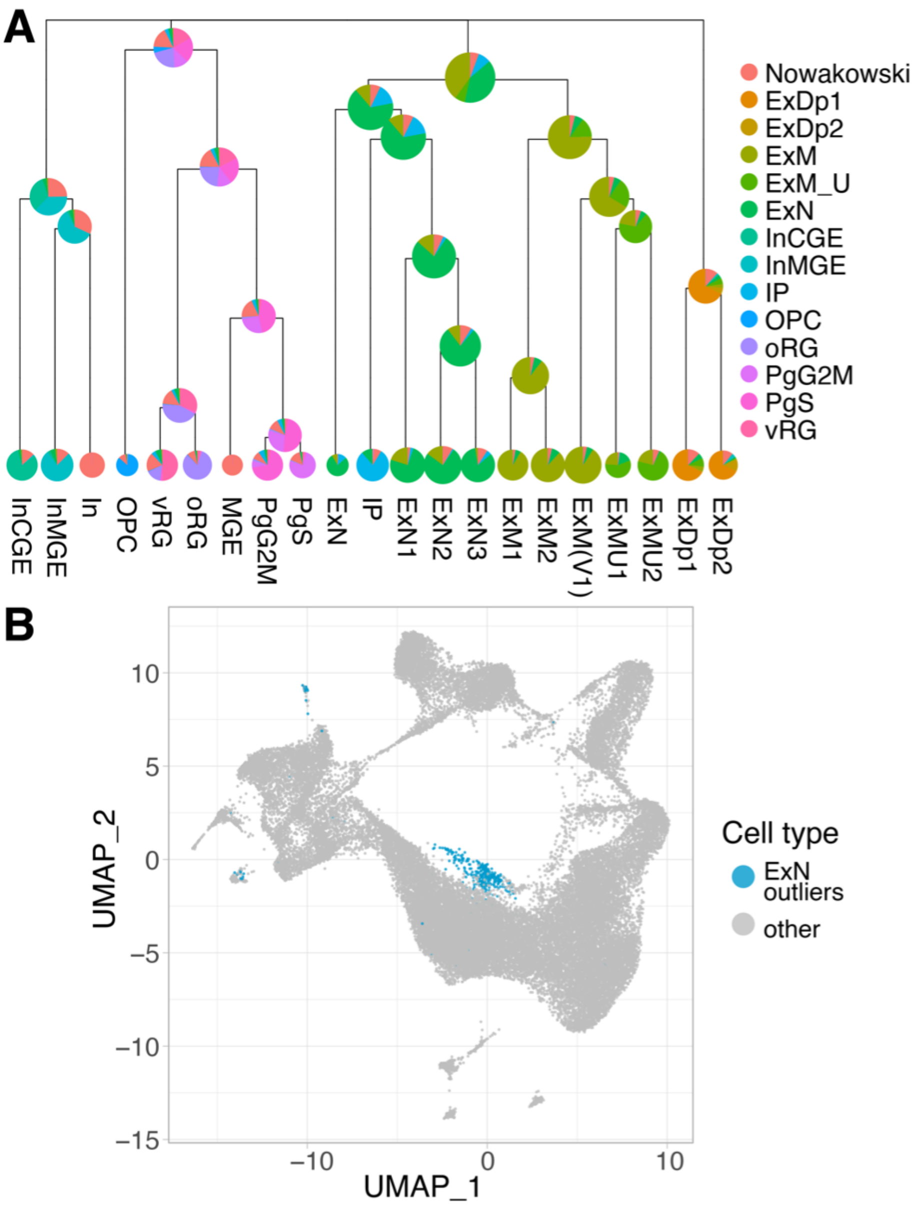


***Supplementary Fig. 8. (a)*** *Results of MRtree applied to the integrated data, resulting in 21 clusters. Nodal labels are spelled out in Figure 5. From the colors of nodes the fraction of cells from the Nowakowski data is apparent.* ***(b)*** *The cluster indicated in blue was considered an outlier and removed from downstream analysis. This small, outlying cluster, originally labeled as ExN by Polioudakis, did not cluster with the other ExN cell types.*

We partitioned this list genes according to whether they were DD or ASD-predominant (posterior probability < 0.01 and > 0.99), which yielded 82 and 36 genes, respectively. For each cluster, we compute the odds ratio from the 2x2 table to determine enrichment. The enrichment analysis was performed for the ASD and the DD-predominant risk genes sets (**Figure 5c**).

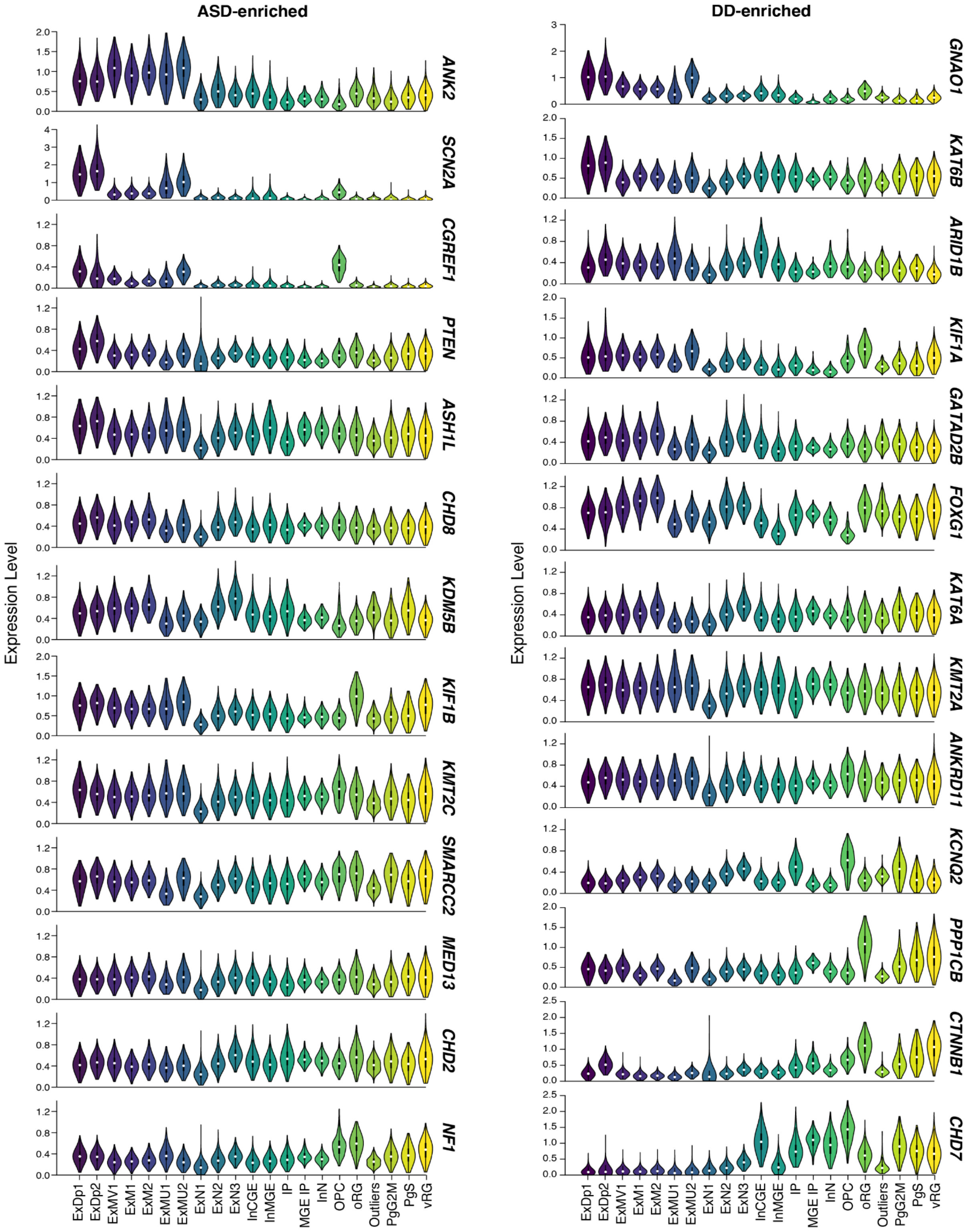


***Supplementary Fig. 9.*** *Top 13 enriched genes from the ASD-predominant and DD-predominant gene lists with expression data in the single-cell dataset. Geres are ordered visually from neuron-enriched (top) to progenitor-enriched (bottom).*

### Supplemental information on Statistical Tests

**Page 2**
Figure 1b,c,e,f: Due to the fact that many individuals harbor more than one inherited variant, we chose to calculate and display the excess of mutations in each inheritance category using a difference in rates (instead of odds ratios) so that all measures are on a comparable scale across *de novo*, case/control, and inherited. For *de novo* variants, the binomial test statistic is the ratio of the *de novo* variants in the affected offspring divided by the rate of *de novo* variants in all offsprings, with the null probability equal to the number of affected offspring over all offsprings. For case/control, the test statistic and null probability are constructed with the equivalent case/control numbers. For the inherited variants, the binomial test statistic is the proportion of transmitted variants, and the null probability is 0.5.

**Page 3**
Comparing odds ratio of deletions and PTVs over first decile of LOEUF: We set the null hypothesis to be that the enrichment of deletions is the same as the enrichment in PTVs. We then randomly generate the number of deletions observed in affected offsprings using the null hypothesis enrichment, multiplied by the base observed rate of mutations in unaffected offsprings to calculate an odds ratio. This process is repeated $10^7$ times, and the p-value is calculated as the number of iterations where the odds ratio exceeded the observed odds ratio.

Logistic regression: For each comparison noted with a logistic regression comparison, we modeled ASD status (or male versus female where appropriate) as the outcome variable, and the two variables of interest as input variables. Regression outputs are then reported.

**Page 4**
Correlation tests: R function `ggpubr::cor.test` was used to evaluate the significance of correlation coefficients where reported.

### Data and Code Availability

The data used in this study are available at:
Repository/DataBank Accession: NHGRI AnVIL
Accession ID: phs000298
Databank URL: https://anvilproject.org/data

Repository/DataBank Accession: Simons Foundation for Autism Research Initiative SFARIbase
Accession ID: SPARK/Regeneron/SPARK_WES_2/
Databank URL: https://www.sfari.org/resource/spark/

The R code used to generate TADA association results will be made available upon reasonable request to authors.
